# Validity Gates Expose Limits of Longitudinal Multimodal Parkinson Disease Prediction

**DOI:** 10.64898/2026.09.23.26363849

**Authors:** Harsh Milind Tirhekar, Priyanshi Yadav, Chandrajit Bajaj

## Abstract

Multimodal Parkinson disease models can improve internally while obscuring why patients differ and whether a gain is deployable. We separated clinical trajectories, dated biological sources, observation process and cohort-relative, age-weighted evidence for dopaminergic, injury/glial, cognitive co-pathology and lysosomal burden across 5,404 cutoffs in 1,058 Parkinson’s Progression Markers Initiative participants. DaT-SPECT and MoCA history produced a small 12-month motor-change gain (Δ*R*^2^ = 0.015; delta MAE=-0.060) that failed prespecified magnitude and strict-calendar criteria. Continuous evidence retained overlapping support but showed no detectable predictive increment beyond raw sources. Source-aware intervals achieved 90.6% simultaneous participant coverage yet failed selective prediction, yielding ABSTAIN. In 8,000 semisynthetic experiments, the gate limited null promotion to 0.35%, detected leakage and blocked 94.75% of calendar reversals; PDBP clinical transport and multimodal eligibility failed. The framework reveals state support, stale or missing evidence, and unjustified prognostic, uncertainty or transport claims.

## 1 Introduction

Longitudinal prediction in Parkinson’s disease (PD) is difficult for reasons that a larger model does not remove. Motor scores fluctuate with examination state and treatment, progression is heterogeneous, and imaging or cognitive measurements are collected at irregular times. Multimodal learning promises to combine these partial views, but an internally improved average error is only one part of the evidence required for a trustworthy forecast [1–3].

For multimodal AI developers, biomarker scientists and clinical researchers, the practical question is therefore not simply which model predicts best. A useful patient-state representation must distinguish the observed clinical phenotype from the biological evidence offered to explain it and from the process by which that evidence was collected. Two participants with similar motor burden can carry different dopaminergic, cognitive, injury-related or lysosomal support, and the same measured value can have different evidential weight when it is current, old, missing or discordant. A point forecast alone hides those distinctions.

Several common design choices can make longitudinal performance look stronger than it is. Repeated visits from one participant can enter both training and testing. A source recorded after the prediction cutoff can leak future information. Missingness and observation age can identify a study protocol or enrollment era rather than disease state. Marginally calibrated intervals can coexist with an uncertainty ranking that fails to identify low-error predictions. Finally, a model that performs internally can fail after a calendar shift or in an independent cohort. These are different failure modes and require different tests.

Recent PD studies have advanced trajectory subtyping, graph-based integration, multimodal progression modeling and modality-specific transport [4–11]. Their diversity also makes a methodological gap visible: performance is often summarized at one internal split, whereas participant dependence, source chronology, uncertainty discrimination and deployment shift are reported unevenly. Reporting guidance such as TRIPOD+AI improves transparency, but a longitudinal multimodal claim also needs an executable rule for when it must stop [12].

We address that gap with a claim-specific validity-gate benchmark for 12-month change in MDS-UPDRS Part III. The benchmark keeps biological values separate from their observation process, groups all cutoffs from one person, and challenges the same prespecified model across participant-disjoint, site-disjoint and strict-calendar evaluations. It then asks a distinct uncertainty question: can source age, availability and cross-source disagreement rank future error better than clinical uncertainty while maintaining simultaneous coverage over each participant’s repeated predictions [13–15]?

Here, mechanism-state severity has a deliberately limited meaning: cohort-relative support for a mechanism family in the available measurements, accompanied by source age and a separate reliability proxy. Within a multi-source coordinate, age weights alter the relative contributions; with only one source, normalization cancels its weight and ageing does not lower the evidence score. A higher coordinate is therefore not a calibrated mechanism probability, clinical severity threshold, causal disease activity, formal biological stage or treatment target. Keeping evidence value and reliability distinct lets the analysis ask whether clinically similar records have different biological support without claiming to explain why the disease arose.

The contribution is methodological rather than a new clinical risk score. First, 5,404 future-only cutoffs from 1,058 PPMI participants quantify the internal value and temporal fragility of dated DaT-SPECT and MoCA history. Second, mutually exclusive participants fit the predictive mean, residual scale, conformal calibration and outer test, preventing calibration reuse. Third, 8,000 semisynthetic experiments test gate behavior under null and stable effects, repeated-participant leakage and calendar reversal without exposing scenario labels to the decision rule. Fourth, an untouched PDBP test and a diagnosis–modality–time intersection audit show why recalibration and archive size do not by themselves establish transport.

Two related preprints from our group construct broad multiscale and posterior-aware PD representations [16, 17]. The present study asks a different question: what evidence must such a representation pass before an internal prediction, an uncertainty estimate or an external claim is promoted? Applied to the present record, the gates retained a small internal information increment, returned ABSTAIN for selective uncertainty, and rejected calendar and external transport claims. A failed gate is retained as a scientific result, and ABSTAIN denotes a research-validity decision rather than a clinical recommendation.

## 2 Results

### 2.1 Similar motor histories can have different evidence support

We begin with two actual records selected without their future outcomes or forecast errors (Figure 1). Cases P1-A and P1-B had motor totals of 10 and 11 and prior slopes of approximately 1.3 and 1.5 points/year. Their current motor summaries were similar, but their supporting records were not. P1-A had contemporaneous putamen SBR and MoCA, with MoCA 19, but no scored injury/glial or lysosomal source. P1-B had MoCA 25 and no observed DaT-SPECT value in the rolling record, but did have NfL and a GCase measurement approximately 14 months old. Treatment documentation also differed: active LEDD was unrecorded for P1-A and recorded for P1-B. Missing exposure does not mean untreated, and these records are not treatment-matched controls.

**Figure 1:**
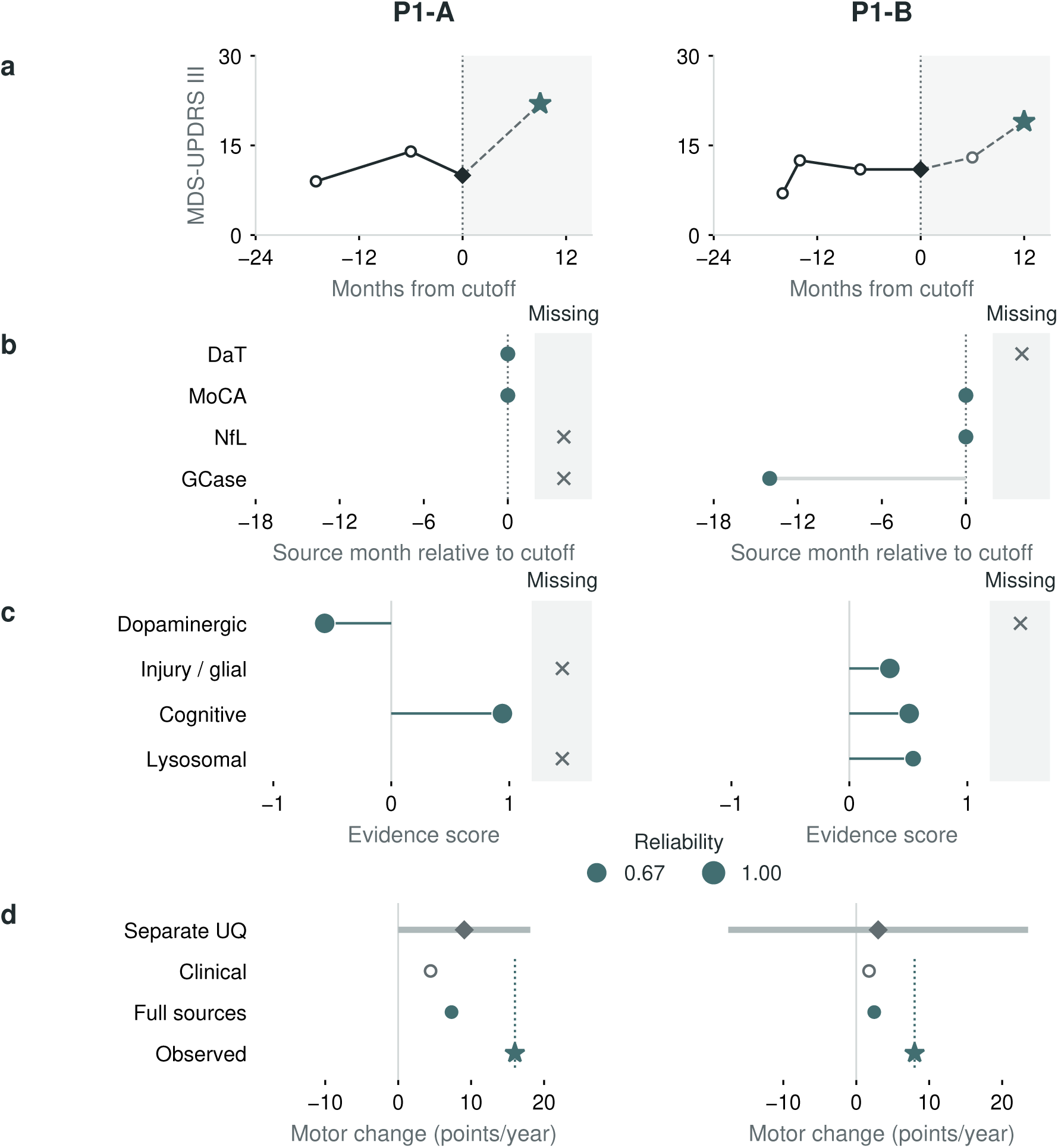
Similar motor histories can rest on different evidence. Columns show outcome-blind Cases P1-A/P1-B. a, Observed monthly motor summaries (circles), cutoff values (diamonds), and selected future summaries (stars); shading and dashed segments identify post-cutoff follow-up. P1-B’s cutoff combines ON and OFF examinations by the primary monthly-median rule, not a single examination. b, Dated sources; zero is the cutoff month, and crosses in the missing-data strip are not zero measurements. c, Training-fold-relative evidence scores; point area represents the separate reliability proxy. Missing coordinates are not normal values; neither quantity is a clinical probability. d, Held-out forecasts and observed annualized change in points/year. The dotted line marks observed change; the gray 90% participant-cluster conformal interval belongs only to the separate uncertainty-model mean (diamond), not the clinical or full-source forecasts. These cases do not override ABSTAIN; Supplementary Section S11 retains all model comparisons.

The clinical-only forecasts were approximately 4.4 and 1.8 annualized motor points; the full source-time forecasts were 7.3 and 2.5. Follow-up motor totals were 22 at nine months for P1-A and 19 at twelve months for P1-B, corresponding to annualized changes of 16 and 8. Neither observed change was used to select the pair. The displayed biological and treatment differences do not establish the cause of their different outcomes.

What integration reveals here is the support behind a patient-state description: which measurements exist, which are old or absent, and which fitted model used them. The two cognitive evidence scores are supported by MoCA, not an adjudicated molecular co-pathology. An identical forecast could therefore have very different evidential foundations. This distinction matters when reviewing a model for monitoring or trial research, but the broad intervals and failed selective-validity gate prevent these examples from becoming individual prognostic or treatment claims.

The examination adds a distinction that the total score hides (Figure 2). P1-A’s paired limb findings were confined to the left body side: finger tapping, hand movements, leg agility and upper-limb rest tremor each scored one. Lower right than left putamen binding (0.71 versus 0.96) was anatomically concordant with that left-body pattern. P1-B had right-predominant findings in both source examinations, but no observed DaT-SPECT value to test anatomical concordance. Its OFF and ON side totals were respectively left/right 0/8 and 1/7. The primary monthly summary preserves medians across those examinations; it does not estimate an ON–OFF treatment response. A motor laterality of -1 in P1-A reflects a denominator of only four paired points, not maximal whole-disease severity. The missing scan in P1-B remains missing rather than becoming a model-imputed brain abnormality.

**Figure 2:**
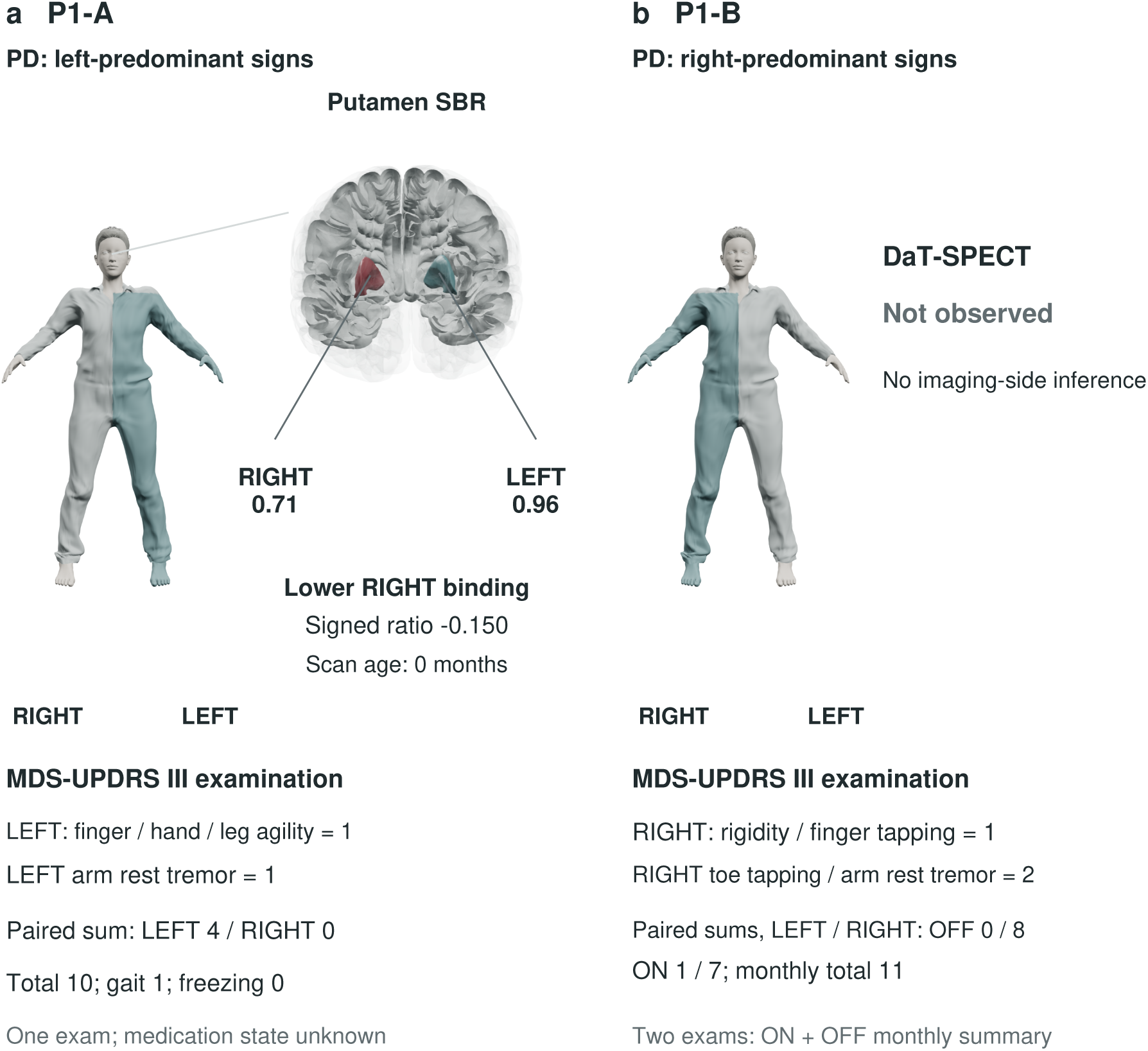
Observed body-side findings and the limits of internal anatomical evidence. a, P1-A has four left-side paired motor points, total MDS-UPDRS III 10, and contemporaneous lower right putamen SBR. Labels give actual scores and hemisphere-specific binding; the reference human and brain are not participant images. b, P1-B has right-predominant signs in both OFF and ON examinations. Its total 11 is a monthly median, and absent DaT-SPECT precludes an imaging-side inference. Highlighted body sides indicate aggregate examination burden, not measured motion or uniform abnormality in every limb item. Scores are ordinal examination findings; no formal tremor-dominant or gait subtype is assigned. Relative lower binding is a within-person contrast, not a normative severity category. Human: MakeHuman/MPFB, CC0; brain: NIH 3D Human Reference Atlas 3DPX-020960, CC BY 4.0.

The relevant anatomical link is the nigrostriatal projection from substantia nigra to striatum within the same hemisphere, interpreted alongside contralateral motor expression through the wider motor circuit. P1-A’s right-putamen/left-body concordance fits this reference organization; it does not measure a direct putamen-to-limb connection or the severity of an entire pathway. The motor and associative loops in Supplementary Figure S1 distinguish this localized imaging support from MoCA and fluid markers, which cannot identify a damaged circuit in either record [18].

### 2.2 From patient records to claim-specific validity tests

The cohort-level results follow the sequence in Figure 3: chronology, internal added value, temporal stress, selective uncertainty, semisynthetic gate behavior and external transport. Each stage can strengthen, narrow or stop a claim.

**Figure 3:**
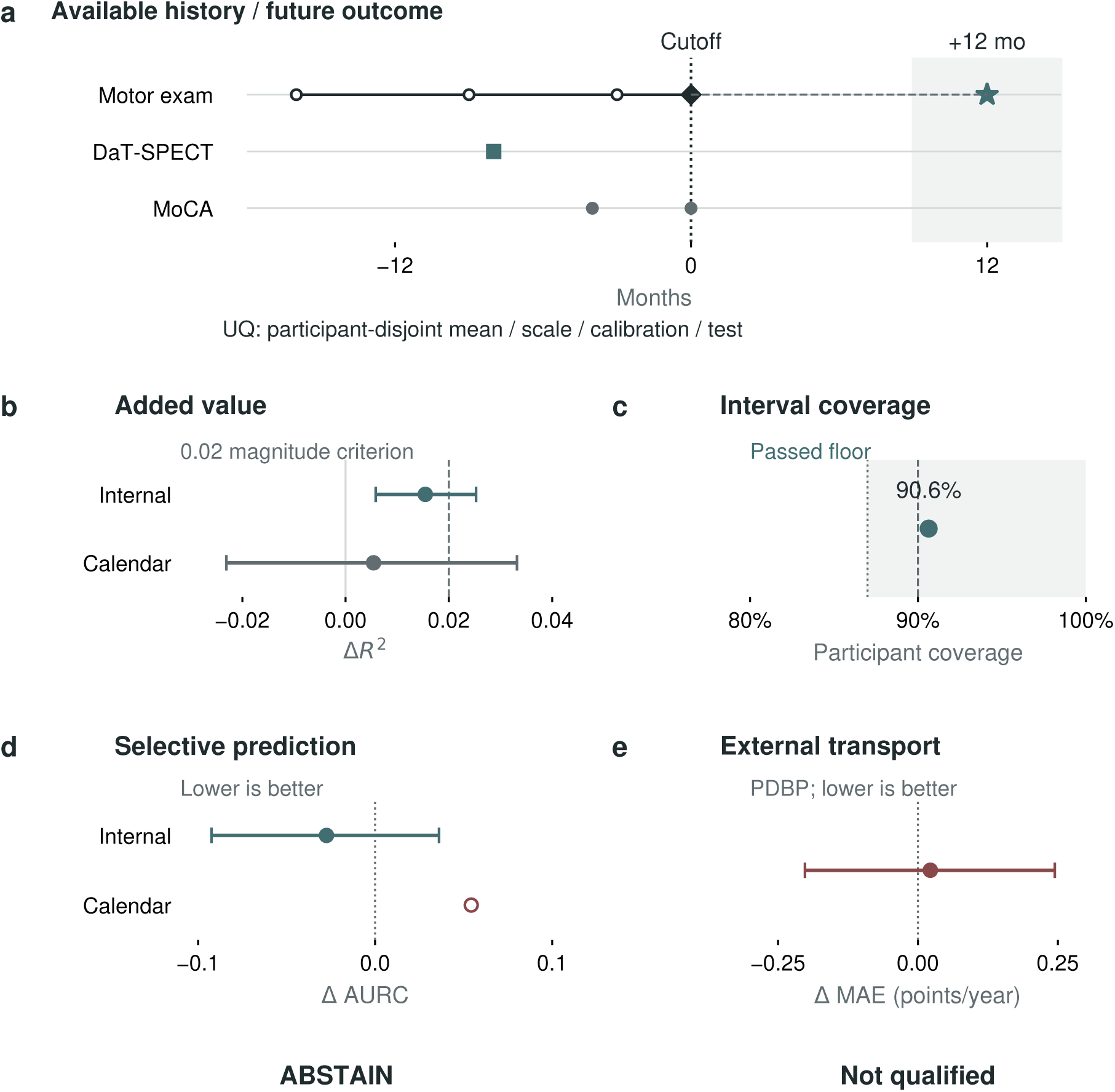
Different validity questions produce different verdicts. a, Schematic cutoff and future 9–15-month endpoint window; circles denote motor/cognitive observations, a square the scan, a diamond the cutoff, and a star the future motor endpoint. This is a design schematic, not a patient history. b, Full-source minus clinical Δ*R*^2^, internally and under strict-calendar deployment, with paired 95% participant-bootstrap intervals; the dashed 0.02 line is the magnitude criterion. c, Internal simultaneous participant coverage is 90.6%; dashed and dotted lines mark nominal 90% and the 87% floor. d, Source-aware minus clinical uncertainty area under the risk–coverage curve (AURC): negative favors source awareness. The internal 95% interval crosses zero; the calendar point reverses, with no interval estimated for that displayed contrast. e, Recalibrated six-variable clinical transport to 156 PDBP participants versus their calibration-mean reference: positive delta MAE is unfavorable. It is not a multimodal external test. Adequate coverage does not override failed selective discrimination or transport.

**Figure 4:**
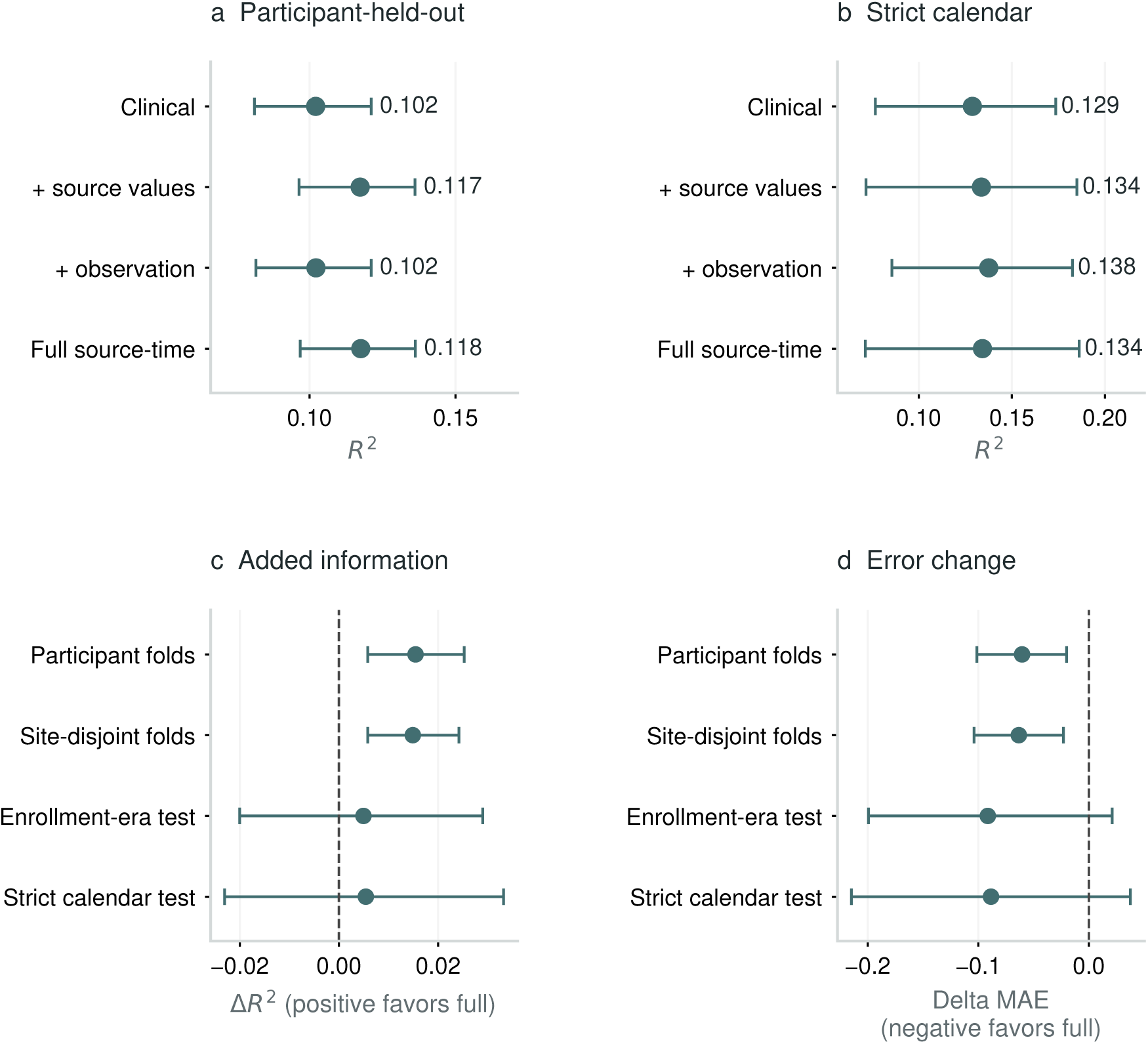
Information-block performance across progressively stricter validation. a, Pooled participant-fold *R*^2^ and 95% participant-cluster confidence intervals for the four information blocks. b, Strict deployment performance after training only on outcomes completed before 2020 and testing later enrollees without refitting. c, Full-source minus clinical Δ*R*^2^ across participant, site, enrollment-era and strict calendar evaluations. Positive values favor the full model. d, Corresponding delta MAE; negative values favor the full model. The figure follows the same source-aware model from an internally detectable increment to an uncertain calendar-era advantage, showing why participant separation alone is not a deployment test.

Table 1 summarizes the resulting validation decisions. A passed criterion permits only the inference in its final column; it cannot substitute for a failed criterion elsewhere in the sequence.

**Table 1:** Claim-specific validity gates and allowed inferences.

| Claim | Evaluation set | Required challenge | Status | Allowed inference |
| --- | --- | --- | --- | --- |
| Mean prediction | PPMI, 1,058 people; 5,404 cutoffs | Participant and site separation; effect-size threshold | Below promotion threshold | Dated sources add a small internal increment below promotion magnitude |
| Calendar deployment | Earlier PPMI to 442 later enrollees | Training outcomes completed before 2020 | Fail | Internal added value is not stable across enrollment era |
| Selective uncertainty | Same PPMI rolling cohort | Separate participants for mean, scale, calibration and test; cluster coverage | ABSTAIN | Coverage passes, but selective discrimination does not |
| Gate validity | 8,000 semisynthetic experiments | Null, stable, leakage and calendar-shift conditions | Protocol behavior supported | Low null promotion and useful power; residual shift failures remain visible |
| Clinical transport | PPMI 1,256 to PDBP 156 test | Prespecified external test and cohort-mean comparison | Fail | Recalibration does not establish transport |
| Imaging feasibility | Local PDBP export | Diagnosis–modality–time intersection | Fail | Files are present, but no eligible idiopathic-PD imaging cohort exists |

**Table 2:** Compact audit of the enrollment-era shift. Proportions use all rolling cutoffs; prior-visit count uses one earliest eligible cutoff per participant. SMD is later minus earlier.

| Variable | Through 2019 | 2020 onward | Shift | Interpretation |
| --- | --- | --- | --- | --- |
| Prior visit count | 4.88 | 3.17 | SMD -1.91 | Observed-history and protocol shift |
| MoCA slope available | 75.3% | 42.0% | -33.3 points | Observation-process shift |
| Active LEDD available | 93.3% | 76.5% | -16.8 points | Treatment-record shift |
| DaT level available | 97.0% | 98.1% | +1.1 points | Stable imaging availability |
| Annualized motor change | 1.31 | 0.34 | SMD -0.11 | Endpoint-distribution shift |

**Table 3:** From scale to measurement and analytical role. All predictor sources are dated at or before the cutoff. The evidence-coordinate analysis is distinct from the primary clinical-plus-source benchmark.

| Scale / information | Measurement | Role and chronology | Interpretation limit |
| --- | --- | --- | --- |
| Clinical motor | Total score, laterality, gait, freezing | Reference: current values and prior slopes | Motor expression, not molecular cause |
| Regional imaging | Putamen SBR, signed asymmetry | Dated values and prior SBR slope; dopaminergic evidence coordinate | Not direct dopamine release or circuit dynamics |
| Clinical cognition | MoCA | Dated value and prior slope; cognitive evidence coordinate | Not a specific co-pathology diagnosis |
| Fluid biomarkers | Assay-specific NFL, GFAP, amyloid/tau and GCase | Pre-cutoff support for the separate evidence-coordinate comparison | Proxies, not calibrated pathway activity |
| Treatment context | Active LEDD; examination state | Dated covariates and state sensitivities | Not causal treatment response |
| Observation process | Source age, availability, visit density | Separate predictor block and uncertainty inputs | May encode study era rather than biology |

### 2.3 A date-resolved cohort separates people from repeated opportunities to predict

The current PPMI release yielded 25,118 dated clinical patient-months from 4,773 participants, including 1,602 participants classified as PD. Of these, 1,256 had an eligible baseline-to-12-month motor outcome, 1,036 had an eligible 24-month outcome, and 1,348 had at least three dated motor observations over at least 12 months. The primary rolling-origin benchmark was stricter. Every cutoff required at least three prior clinical observations, at least 12 months of observed history, and a future MDS-UPDRS Part III assessment nearest 12 months within a plus or minus 3-month tolerance. This produced 5,404 cutoffs from 1,058 PD participants.

The sample size for generalization is therefore 1,058 people, not 5,404 independent people. All cutoffs from one participant remained in one outer fold. Bootstrap draws sampled participants and retained every eligible cutoff from each sampled participant. Source values observed after a cutoff were never allowed into that cutoff’s feature vector.

The 1,058 participants were also divided by an enrollment-era proxy defined before modeling: the earliest dated PD motor observation in the release. Participants first observed through 2019 formed the development era (616 participants; 4,664 cutoffs), and participants first observed from 2020 onward formed the untouched temporal cohort (442 participants; 740 cutoffs). The mean annualized motor change was 1.31 points per year (SD 9.39) in the earlier era and 0.34 (SD 8.95) in the later era. DaT-SPECT level coverage was high in both eras (97.0% and 98.1% of cutoffs), but MoCA slope coverage fell from 75.3% to 42.0%, and active LEDD coverage fell from 93.3% to 76.5%. The temporal test therefore changes both participants and observation patterns.

At the earliest eligible cutoff, participants had mean age 63.8 years (SD 9.7), 407 (38.5%) were recorded female, median diagnosis-derived disease duration was 1.8 years (interquartile range 1.4-2.8), and mean MDS-UPDRS Part III was 23.5 (SD 10.9). Of 792 participants with recorded exposure, 775 (97.9%) had active LEDD; exposure was unavailable for 266. Recorded race was White for 1,011 (95.6%), and 53 (5.0%) had Hispanic or Latino ethnicity. PPMI race categories permit multiple selections. Age was unavailable for three participants. The small non-White cells precluded reliable race-stratified performance assessment; complete aggregate characteristics are reported in Supplementary Section S2.

PDBP clinical transport and the PDBP image-feasibility audit were separate analysis sets and were never added to the PPMI count. This layered design is more than cohort bookkeeping: it prevents repeated visits, different diagnoses and different scientific questions from being collapsed into one apparently large but invalid sample.

### 2.4 Multimodal history adds information internally but little clinical value

The clinical reference contained current motor total, signed clinical laterality, gait, freezing, active LEDD, number and span of prior visits, pre-cutoff motor and laterality slopes, age, recorded sex code and diagnosis-derived disease duration. The dated source-value block added current putamen mean SBR, signed putamen asymmetry, pre-cutoff putamen SBR slope, current MoCA and pre-cutoff MoCA slope. The observation-process block added time since the most recent DaT-SPECT and MoCA measurements and explicit indicators of whether each level and slope was available. The full model contained all three blocks.

Across prespecified five-fold participant splits, the clinical reference achieved *R*^2^ 0.102 (95% participant-bootstrap CI 0.081 to 0.121), MAE 6.766 and RMSE 8.846. Adding dated source values increased *R*^2^ to 0.117 and reduced MAE to 6.706. The paired contrast was Δ*R*^2^ 0.015 (95% CI 0.006 to 0.025) and delta MAE -0.061 (-0.100 to -0.021). Observation-process variables alone did not improve the internal reference (Δ*R*^2^ 0.000, -0.003 to 0.003; delta MAE -0.005, -0.017 to 0.007). The full source-time model achieved *R*^2^ 0.118 and MAE 6.706, with Δ*R*^2^ 0.015 (0.006 to 0.025) and delta MAE -0.060 (-0.101 to -0.020).

These results identify where the internal increment resides. It is carried by dated DaT-SPECT and MoCA levels and slopes, not by source-age or availability variables in the pooled participant-fold analysis. The improvement is statistically stable but clinically very small: a 0.060-point reduction in MAE on MDS-UPDRS Part III does not establish usefulness for patient management or trial enrichment. For scale only, anchor-based estimates of meaningful OFF-state motor worsening are 4–6 MDS-UPDRS Part III points, with clinical consensus at 5 points [19]; that threshold concerns within-person worsening rather than prediction error and is not a utility cutoff for this mixed-state observational benchmark. Even so, it makes the difference in magnitude unmistakable. The internal result also does not clear the prespecified added-value criterion of Δ*R*^2^ at least 0.02 with an interval above zero.

The internal signal survived outcome-blind alternatives. Non-stratified GroupKFold produced Δ*R*^2^ 0.018 (95% CI 0.008 to 0.028) and delta MAE -0.068 (-0.108 to -0.025). Equal participant weighting and earliest- or latest-cutoff analyses also retained positive increments (Supplementary Section S5). When acquisition site defined the held-out unit, the full model achieved *R*^2^ 0.114 versus 0.099 for the clinical reference across 951 participants from 52 sites (Δ*R*^2^ 0.015, 0.006 to 0.024; delta MAE -0.063, -0.104 to -0.023). These tests support a small within-PPMI information increment; they do not establish external transport or clinical importance.

Endpoint-noise analyses reached the same bounded conclusion. For the primary held-out predictions, the full-minus-clinical change in median absolute error was -0.036 points (95% participant-cluster bootstrap CI -0.130 to 0.086). We then winsorized the training outcome at its fold-specific 1st and 99th percentiles, refitted every imputation, scaling and ridge step, and bounded each held-out outcome and prediction using only the corresponding training limits. The internal Δ*R*^2^ was 0.0157 (0.0060 to 0.0258), delta MAE was -0.061 (-0.099 to -0.021), and delta median absolute error was -0.024 (-0.125 to 0.080). Under enrollment-era transport, winsorized Δ*R*^2^ was 0.0084 (-0.0151 to 0.0328) and delta MAE was -0.100 (-0.206 to 0.004). Extreme motor changes therefore did not create the internal signal, but robust endpoint handling did not restore calendar promotion.

Motor examination state was evaluated separately because MDS-UPDRS Part III depends on dopaminergic treatment. Adding explicit cutoff ON/OFF indicators to both models produced Δ*R*^2^ 0.019 (95% CI 0.009 to 0.029) and delta MAE -0.069 (-0.113 to -0.027). Direction was retained among ON-to-ON observations (1,348 cutoffs; Δ*R*^2^ 0.017, 0.001 to 0.034) and was imprecise among the much smaller OFF-to-OFF subset (163 cutoffs; Δ*R*^2^ 0.029, -0.015 to 0.076). Outcome exam state defined sensitivity strata only and was never a predictor. These observational checks do not estimate medication effects.

The internal signal is therefore detectable and repeatable, but too small to support a clinical claim. Its value is to motivate a harder question: does the same increment remain credible when the study calendar changes?

### 2.5 The internal gain weakens when the calendar moves

All four fixed ridge models were next fit using only the 616 earlier-era participants and evaluated without refitting or recalibration in the 442 post-2020 participants. The clinical reference achieved *R*^2^ 0.131 (95% CI 0.078 to 0.176), MAE 6.542 and calibration slope 1.055. Dated source values achieved *R*^2^ 0.135 and MAE 6.462, but their paired improvement was uncertain (Δ*R*^2^ 0.004, -0.019 to 0.026; delta MAE -0.080, -0.180 to 0.023).

Observation-process variables alone produced a small interval-supported change in this temporal cohort (*R*^2^ 0.138; Δ*R*^2^ 0.008, 0.001 to 0.015; delta MAE -0.050, -0.080 to -0.020). In contrast, the full source-time model did not provide an interval-supported advantage: *R*^2^ 0.136, MAE 6.451, Δ*R*^2^ 0.005 (-0.020 to 0.029) and delta MAE -0.091 (-0.199 to 0.021).

This pattern is informative. A measurement schedule can carry cohort-era information even when the corresponding biological values do not transport. The marked reduction in MoCA slope and LEDD coverage in the later era supports that interpretation. The result does not justify using observation frequency as a clinical biomarker. It shows why value, age and availability should be reported separately and why internal multimodal gains require temporal or external challenge.

Because enrollment-era separation alone allows later outcomes from legacy participants to enter development, a stricter sensitivity excluded every training row whose outcome occurred on or after 1 January 2020. This left 580 earlier participants and 3,575 training cutoffs; the same 442 later enrollees remained untouched. The full model again gave an uncertain increment over the clinical reference (Δ*R*^2^ 0.005, 95% CI -0.023 to 0.033; delta MAE -0.089, -0.215 to 0.038). Observation-process variables retained a small association (Δ*R*^2^ 0.009, 0.001 to 0.016), whereas dated source values did not.

The eras were not exchangeable. A five-fold logistic model using one earliest predictor history per participant distinguished later from earlier enrollees with AUC 0.978 (95% participant-bootstrap CI 0.969 to 0.987). The largest shift was the amount of available clinical history (standardized mean difference -1.91); disease duration, age, DaT-SPECT level, MoCA and source-slope availability also shifted. This domain-classification result is a dataset-shift diagnostic, not a disease classifier.

A secondary subgroup audit found no interval-supported temporal increment within any broad age or recorded-sex stratum. These descriptive analyses neither tested interactions nor established subgroup fairness; complete counts and estimates are reported in Supplementary Section S6.

The calendar challenge changes the verdict. Information that was stable across participants and sites did not become a demonstrable deployment advantage in a later acquisition era.

### 2.6 Source-aware uncertainty is calibrated but does not clear selective-prediction gates

Average error can hide where a model is unreliable. We therefore represented each source as a dated record containing its value, observation time, age, availability, provenance and a deterministic reliability coordinate. DaT-SPECT and MoCA reliability decayed with source age; clinical-history reliability increased with longitudinal density. Three fixed disagreement coordinates compared normalized motor severity with dopaminergic deficit, motor with cognitive deficit, and motor with dopaminergic velocity. These variables were permitted to predict uncertainty, not the future endpoint directly.

The evaluation was nested. For every outer test fold, predictive-mean residuals were generated inside the remaining participants. A participant-disjoint subset fitted the residual-risk model, a separate subset calibrated a nominal 90% interval, and the untouched outer participants supplied risk–coverage and calibration metrics. The same transparent full-source ridge predictor was used for all uncertainty comparisons. Clinical uncertainty used clinical trajectory and context; generic uncertainty added raw dated sources; source-aware uncertainty further added reliability and disagreement. Thus any uncertainty difference arose from the uncertainty representation rather than a different mean predictor.

The full mean predictor achieved participant-held-out *R*^2^ 0.103 and MAE 6.772 under this inverse-participant-weighted nested design. At 90%, 80%, 70% and 50% retention, source-aware MAE was 6.614, 6.478, 6.396 and 6.129 points, respectively. Clinical uncertainty reached 6.637, 6.526, 6.391 and 6.207 at the same retentions. Integrated from 50% to 100% retention, AURC was 6.465 for clinical, 6.449 for generic multimodal and 6.437 for source-aware uncertainty. The source-aware-minus-clinical difference was -0.027 (95% participant-bootstrap CI -0.092 to 0.036). The first promotion requirement therefore failed.

Conformal calibration did not rescue discrimination. Calibration used each participant’s maximum normalized residual so that the guarantee addressed a complete repeated-prediction trajectory rather than an exchangeable row. Source-aware intervals covered 97.7% of rows and all eligible rows simultaneously for 90.6% of participants, with mean width 47.82 motor-change points and uncertainty–error Spearman correlation 0.115. That width was approximately 5.1 times the observed endpoint standard deviation of 9.34 points, underscoring the absence of individual prognostic precision despite nominal coverage. The corresponding clinical intervals achieved 97.9% row and 90.9% simultaneous participant coverage with width 49.24. This conservative row coverage is expected when the calibration target is participant-simultaneous. Availability of at least one historical value was also highly imbalanced: 5,245 of 5,404 cutoffs (97.1%) had both a DaT-SPECT and MoCA value, 150 had MoCA only, 7 had DaT-SPECT only and 2 had neither. These strata encode value availability rather than the lower availability of longitudinal slopes reported above; the rare missing-source patterns cannot support independent calibration claims. Under site-disjoint evaluation, source-aware minus clinical AURC was -0.030; under strict 2020-onward deployment it reversed to 0.054 in the unfavorable direction. The locked output was therefore ABSTAIN: participant trajectories were covered, but the uncertainty representation did not identify a transportable low-risk subset (Figure 5a–c).

**Figure 5:**
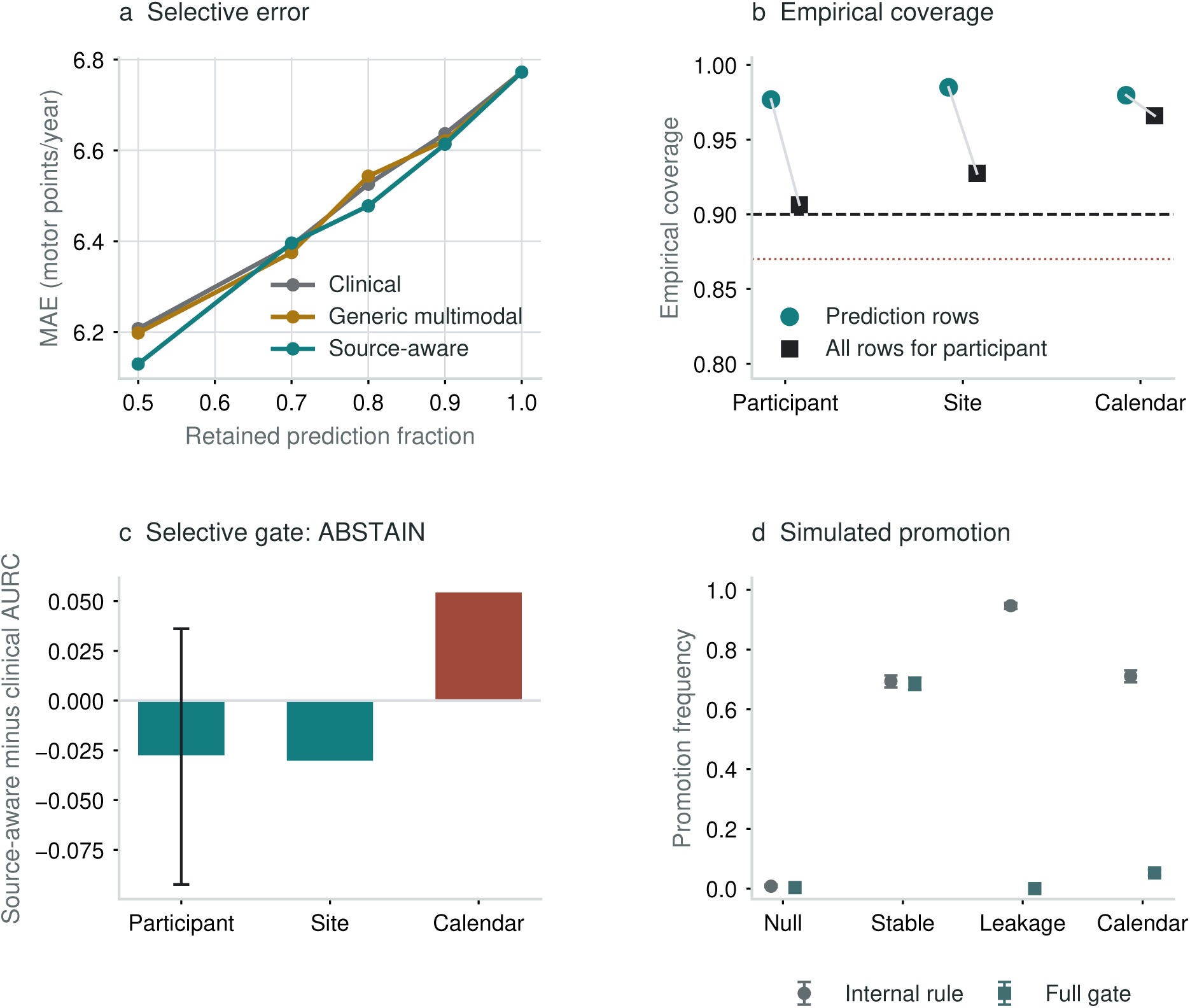
Calibrated uncertainty does not imply selective or transport validity. a, MAE after retaining predictions from lowest to highest uncertainty for clinical, generic multimodal and source-aware uncertainty in 5,404 participant-held-out cutoffs. b, Row-wise and participant-simultaneous coverage of source-aware 90% cluster split-conformal intervals under participant, site-disjoint and strict-calendar challenges; dashed black and dotted gray lines mark nominal 0.90 coverage and the prespecified 0.87 floor. c, Source-aware-minus-clinical AURC over 50%–100% retention. Negative values favor source awareness; the participant error bar is the two-sided 95% participant-bootstrap interval. The internal interval includes zero and the calendar contrast reverses, yielding ABSTAIN. d, Promotion frequencies with binomial 95% intervals across 2,000 semisynthetic replicates per condition. The complete gate reduces null promotion to 0.35%, blocks all detectable overlap, blocks 94.75% of calendar reversals and retains 68.6% stable-effect power. Simulations test the protocol, not disease biology.

**Figure 6:**
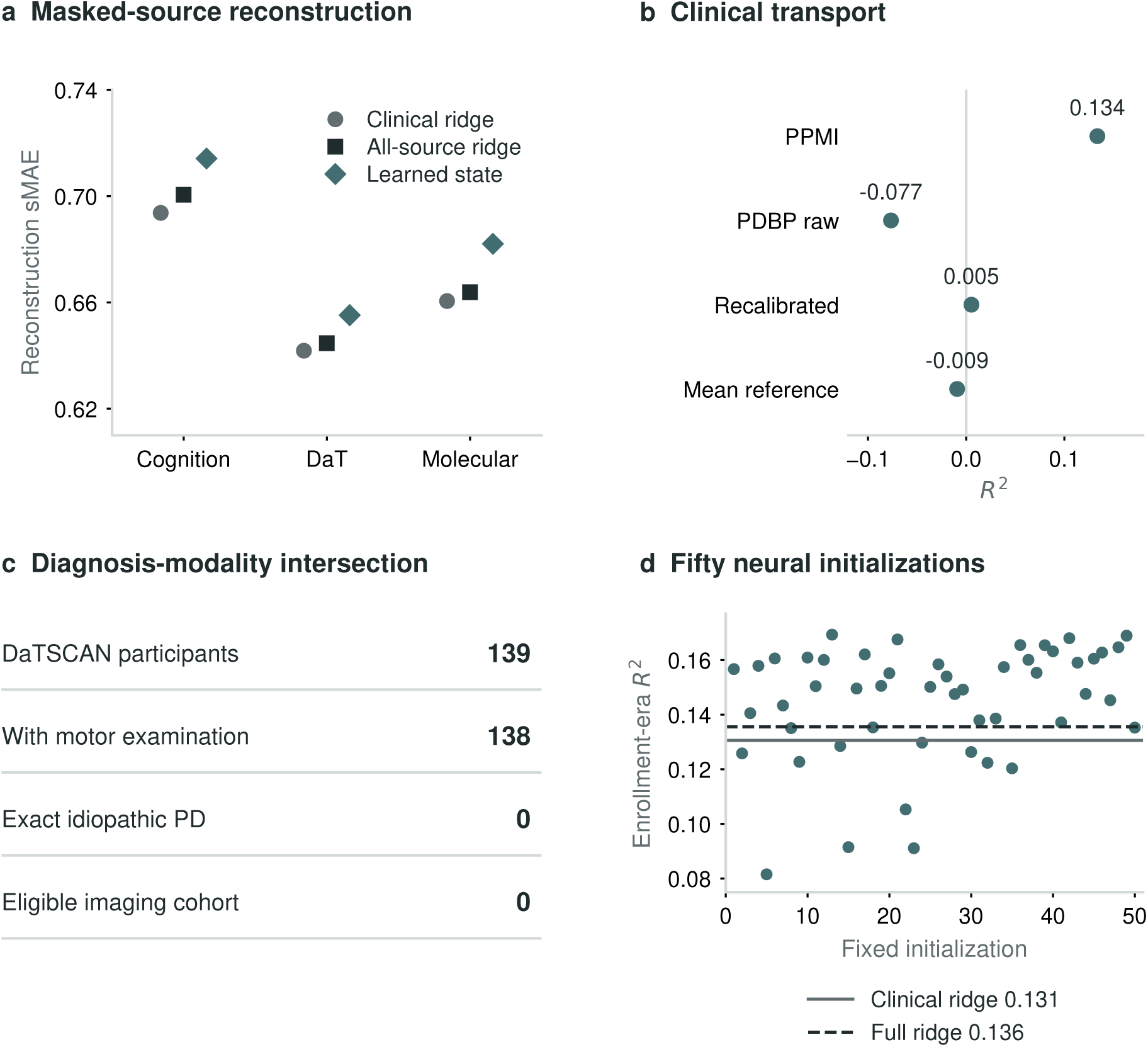
Model-family and transport stress tests. a, Whole-source reconstruction standardized mean absolute error (sMAE): absolute error divided by the fold-local training target standard deviation, averaged within target and then across targets. Symbols are model point estimates, not uncertainty intervals; lower is better. b, Internal PPMI and external PDBP *R*^2^ point estimates for a harmonized clinical-motor model. Recalibration does not beat the calibration-mean reference on mean absolute error (MAE). c, Present DaTSCAN records, available motor examinations and diagnosis eligibility are separate requirements; no eligible idiopathic-PD imaging cohort remains. All 2,442 unique catalog image references were present. d, All fifty fixed neural initializations in the untouched post-2020 enrollment cohort; solid and dashed lines show clinical and full-source ridge references. Thirty-six runs exceeded full ridge *R*^2^, but only 24 improved MAE. No seed was selected or ensembled. Panels show distinct tests, not evidence that model complexity generally fails.

#### 2.6.1 Why coverage did not become selective utility

Five observations explain the failed uncertainty gate. The predictive mean changed little when multimodal history was added, leaving limited error structure for a residual model to recover. Source age, availability and cross-source disagreement weakly ranked future error. Participant-maximum conformal scores were intentionally conservative because one interval failure anywhere in a person’s trajectory counted against simultaneous coverage. Most importantly, source-aware AURC reversed direction across the calendar boundary. Coverage therefore answers whether intervals contain outcomes under the stated exchangeability design; it does not show that uncertainty can identify a useful subset of lower-risk forecasts. Passing coverage while failing discrimination is precisely why the protocol returns ABSTAIN rather than relabelling calibration as triage utility.

We next asked whether the gate, rather than the failed real-data model, behaved predictably. A semisynthetic audit generated 2,000 independent residual-scale-fit, calibration, internal-test and calendar-test cohorts for each of four conditions, for 8,000 experiments overall. Under a null source effect, a naive internal decision promoted 0.8% of replicates and the complete gate promoted 0.35%. Under a stable source effect, promotion power was 68.6%. A measurable participant fingerprint caused naive promotion in 94.7% of leakage replicates, but direct participant-set intersection detected every constructed overlap and blocked all final promotions. Under calendar reversal, 71.1% passed the naive internal decision, whereas 5.25% passed all gates; the calendar challenge therefore blocked final promotion in 94.75% of shift replicates, not all of them (Figure 5d). These simulations validate behavior of this implementation under controlled conditions; they do not validate Parkinson disease biology or make the gate an oracle.

### 2.7 Evidence coordinates improve interpretation rather than prediction

The mechanism-state prototype represented available biology as four non-exclusive evidence coordinates: dopaminergic deficit, injury/glial burden, co-pathology/cognitive burden and lysosomal burden. Values were transformed by training-fold empirical percentiles, oriented by fixed biological direction, and exponentially down-weighted with a 24-month observation-age half-life. The coordinates are evidence summaries, not probabilities, formal biological stages or causal mechanisms.

The coordinates answer which measured mechanism family is most strongly supported after accounting for observation age, not which mechanism caused the patient’s PD. Lower putamen SBR supports the dopaminergic coordinate; higher NfL and GFAP support injury/glial burden; amyloid, tau and MoCA contribute co-pathology/cognitive evidence; and lower GCase activity supports lysosomal burden. A high value therefore means stronger relative evidence in this cohort, conditional on what was measured and when. It is not a clinical severity threshold.

On the same prespecified folds, continuous evidence achieved *R*^2^ 0.114 and MAE 6.726, compared with *R*^2^ 0.103 and MAE 6.764 when each cutoff was forced into one dominant state. Preserving overlapping evidence improved *R*^2^ by 0.011 (95% CI 0.002 to 0.019) and MAE by -0.038 (-0.076 to -0.000). Continuous evidence also improved *R*^2^ over the clinical reference by 0.012 (0.003 to 0.020), although its MAE interval touched zero.

However, raw dated source history remained the better and simpler predictor. Adding evidence coordinates to raw source dynamics changed *R*^2^ by -0.000 (95% CI -0.006 to 0.005) and MAE by 0.010 (-0.016 to 0.037). The evidence coordinates therefore serve as a clinician-readable compression and a missing-evidence audit. They show why two records with similar clinical burden need not represent the same evidential state, but they are not a source of additional prognostic information once the original values and trajectories are available.

Their contribution is explanatory rather than predictive: they preserve overlapping biological support, source age and absent evidence without pretending that a more interpretable name creates new information. This matters for research review because an identical forecast supported by current imaging and cognition is not evidentially equivalent to one produced with stale or missing sources. The present scores do not establish which test to order or which therapy to select.

### 2.8 Neither model complexity nor recalibration rescues failed transport

Whole-source masking tested whether a learned partial-view state could reconstruct an entirely missing source. The learned evidence-state model was worse than clinical ridge for cognition, DaT-SPECT and molecular targets, and a calibrated hybrid provided no secure increment. Across three complete five-fold runs, the neural temporal candidate reached *R*^2^ 0.124–0.131 but did not securely outperform transparent source history. Whole-source ablations supplied no evidence of dependence on DaT-SPECT, digital or provenance blocks, while removing molecular inputs improved performance (Supplementary Section S7). The learned candidate was therefore not promoted.

Internal seed consistency did not extend to the enrollment-era challenge. Fifty fixed initializations were trained only in the earlier-era development cohort and evaluated without seed selection or ensembling in the same 442 later-era participants. *R*^2^ ranged from 0.082 to 0.169 (mean 0.145, standard deviation 0.021) and MAE from 6.354 to 6.716. Thirty-six of 50 runs exceeded the full ridge model’s *R*^2^ of 0.136, but only 24 improved its MAE of 6.451. The architecture therefore showed initialization-sensitive temporal performance and was not promoted.

PDBP provided a separate transport challenge. A six-variable clinical-motor model was trained in 1,256 PPMI participants, recalibrated using 77 prespecified PDBP calibration participants, and evaluated once in 156 untouched PDBP participants with a conservative final PD diagnosis. Internal PPMI *R*^2^ was 0.134. Raw PDBP transport gave *R*^2^ -0.077; recalibration improved *R*^2^ to 0.005 and calibration slope to 0.741. MAE remained 5.882, compared with 5.860 for the PDBP calibration-mean reference (delta 0.022, 95% CI -0.202 to 0.245). The no-material-degradation criterion failed. PDBP is therefore a failed clinical transport test, not external validation of the multimodal model.

The PDBP imaging audit separated file presence, diagnosis, modality and timed-outcome eligibility. The local 3.49-GiB outer bundle contained 4,917 members, including 1,230 ZIP members, 876 direct .dcm files and 51 .ima files. All 2,442 unique catalog references, including all 1,030 unique DaTSCAN references, were present. File absence was not the barrier.

The failure occurred at the diagnosis–modality–time intersection, not at motor-schema availability. The DaTSCAN catalog contained 1,045 records from 139 participants; 138 had at least six valid paired MDS-UPDRS III domains, 137 had a complete 11-pair examination and 78 had complete baseline plus 9–15-month examinations. Yet none had an exact neurological-exam diagnosis of idiopathic PD. Of the 53 DaTSCAN participants represented in the separate diagnosis-change form, none had Parkinson’s disease in either its initial or most-recent diagnosis field. Diagnoses were dominated by dementia with Lewy bodies and related cognitive strata. No DaT participant intersected the curated confirmed-PD timed clinical spine. Consequently no PDBP image was processed and no PDBP SBR was estimated. This export cannot validate multimodal imaging transport in idiopathic PD.

Greater model capacity and cohort-specific recalibration changed numerical outputs, but neither repaired the failed transport claim. The PDBP audit further shows that a large archive is not an external validation cohort until diagnosis, modality and time intersect in the same eligible people.

## 3 Discussion

This study tested a simple but often neglected proposition: a longitudinal multimodal model should be judged by the claim it is intended to support, not by one favorable internal metric. The same dated source history was therefore required to survive separate participant, site, calendar, uncertainty and external-transport challenges. The resulting contribution is not a winning architecture. It is an executable benchmark that makes failure visible before an internal association is promoted as a deployable forecast.

The prognostic result illustrates why that separation is necessary. Dated DaT-SPECT and MoCA history contributed information beyond clinical trajectory across participant- and site-disjoint evaluations. The increment was real in the statistical sense, but small: Δ*R*^2^ was 0.015 and MAE fell by only 0.060 MDS-UPDRS Part III points. Published minimally important within-person changes are -3.25 points for improvement and 4.63 points for worsening [20]. Prediction error and clinical change are different estimands and cannot be equated, but the orders-of-magnitude contrast makes the practical limitation unmistakable. Thousands of repeated cutoffs can narrow an interval; they cannot transform a negligible error reduction into clinical utility.

We therefore do not claim individualized prognosis, trial enrichment or decision support. The internal estimate is an information-value result whose clinical and deployment boundaries are part of the finding.

The temporal challenge then changed the scientific verdict. The full source-time increment was uncertain when models trained on pre-2020 outcomes met later enrollees. Meanwhile, observation-process variables acquired a small association and predictor history distinguished enrollment eras with AUC 0.978. The model could therefore learn not only from what was measured, but from when and how the study chose to measure it. Missingness and measurement age are useful provenance, yet they can encode protocol and care processes rather than disease. Reporting biological values, source age and availability as separate blocks makes that distinction visible.

The selective-UQ result makes the same point more sharply. Participant-cluster calibration protected approximately 90% of complete participant trajectories, which necessarily produced conservative coverage for individual rows. Yet source-aware ranking of low- and high-error cutoffs was not demonstrably better than clinical uncertainty and reversed in the calendar challenge. Simultaneous coverage asks whether all eligible forecasts for a participant are protected at the advertised frequency; selective prediction asks whether uncertainty can identify a subset with lower risk. One can hold without the other. Reporting only row-wise coverage would have made the uncertainty model appear successful while hiding its failed intended use. The locked ABSTAIN verdict prevents that substitution.

The semisynthetic benchmark provides a positive methodological result without changing the real-data verdict. Under a stable source–error relation, the complete gate retained 68.6% promotion power. It reduced null promotion to 0.35%, detected and blocked every constructed repeated-participant leakage case, and blocked 94.75% of calendar-reversal cases. The remaining 5.25% shift promotions are important: a finite calendar test is a strong safeguard, not an oracle. These rates varied over sample size, visit count, participant heterogeneity, missingness and effect strength. They characterize this implementation rather than establish universal thresholds.

Continuous evidence coordinates were useful for a different reason. They preserved overlapping dopaminergic, injury/glial, cognitive co-pathology and lysosomal support better than forcing each person into one hard subtype. A scalar forecast says how much change is expected; the evidence composition says which measured source families support the current state and how reliable that support is. Thus, two participants can have similar motor trajectories and similar forecasts while differing in biological support, source freshness and unresolved missingness. Yet the coordinates added no signal after the raw dated sources were present. Their role is interpretive rather than prognostic. This distinction also separates the present study from our prior precision-stratification and posterior-aware phenotyping preprints [16, 17]. Those studies construct patient states; the present work tests when such states deserve biological, prognostic or decision-level trust. An interpretable name is valuable, but it does not manufacture information.

That distinction has clinical significance even though the present model has no clinical utility. Equal point predictions should not be treated as equally well-supported patient states, and a calibrated interval should not be treated as evidence that uncertainty can identify safer patients. The framework makes those differences reviewable before a model is used for trial stratification, monitoring or decision support. It does not itself recommend a scan, assign a biological subtype or select therapy.

Model complexity did not bypass these boundaries. Learned partial-view states reconstructed missing modalities worse than transparent ridge baselines; whole-source ablations did not demonstrate dependence on the added sources; and neural temporal performance varied across 50 untouched initializations. PDBP recalibration improved a poor raw transport result but did not beat a cohort-specific mean in MAE. A plausible architecture, stable internal seeds and acceptable calibration are not substitutes for source dependence, added value and transport. Retaining these null and failed-gate results counters the common tendency to report only the most favorable split or model.

The PDBP image audit reveals an equally important data lesson. The image files were present; the intended diagnosis–modality–time intersection was not. Large tables can describe different subsets, and a large archive can still contain no eligible external validation cohort. Multi-source inventories must be joined at participant, diagnosis, visit and outcome before sample size is interpreted. Otherwise, apparent scale conceals an empty intersection.

These findings sit alongside, rather than oppose, recent modality-specific advances. Wearable studies have shown reproducible longitudinal sensitivity, and external magnetoencephalography analysis explained meaningful progression variance in its own task [10, 11]. Cross-cohort plasma work, however, found that fusion did not outperform the transporting proteomic view and that a derived severity index lacked an independent longitudinal association [9]. The useful question is not whether any modality predicts any Parkinson disease endpoint. It is whether the claimed increment survives the validation regime appropriate to that modality, endpoint and intended use.

Several limitations define the reach of our conclusions. PPMI is an observational research cohort with non-random assessment schedules, treatment exposure and substantial motor-score variability. Active LEDD and recorded exam state do not eliminate confounding by indication, and exam state was unknown at many visits. Enrollment era is an operational proxy based on the earliest dated PD motor observation, not an adjudicated recruitment date. The calendar and site tests remain internal to PPMI, and site metadata were unavailable for 107 benchmark participants. The population was 95.6% recorded White and 61.5% male; secondary age and recorded-sex summaries were descriptive, sparse non-White cells precluded responsible race-stratified evaluation, and demographic fairness or transport cannot be claimed.

Other limitations are source-specific. The outcome-stratified participant folds balanced internal difficulty, although non-stratified GroupKFold gave the same conclusion. Selective uncertainty was evaluated in PPMI only; site and calendar tests are internal challenges, not independent external validation, and source-availability patterns were dominated by records containing both DaT-SPECT and MoCA. The four-way participant partition reduces the data available to each estimator and produces conservative, wide simultaneous intervals. The semisynthetic benchmark models generic error, leakage and shift rather than PD pathophysiology, and its detectable overlap case does not cover every subtler form of leakage. PDBP transport used six harmonized clinical variables rather than the full multimodal state, while the local imaging export lacked an eligible idiopathic-PD intersection. None of the present analyses establishes individualized prognosis, clinical utility, causal mechanism or subgroup fairness.

The next decisive experiment is an untouched external longitudinal cohort with harmonized clinical outcome timing and enough dated source overlap to evaluate the same mean-prediction and uncertainty analyses without redefining predictors after inspection. Until then, four practices are supported: keep every predictor before its cutoff; split, calibrate and resample by participant; report row and simultaneous participant coverage separately; and challenge internal gains across time and cohort before promotion. Multimodal evidence is not weakened by these limits. Its scientific meaning becomes clearer because the claim is allowed to travel only as far as the validation supports.

A patient-specific digital twin would require a longer validation ladder: reproducible state estimation, externally transported forecasting, causal treatment-response estimation, intervention simulation or optimal control, and prospective evidence that the resulting policy improves outcomes. The present work evaluates qualification of state and forecast claims only, and several gates fail. It therefore provides safeguards for future therapeutic digital-twin research, not evidence that virtual intervention or treatment optimization is currently possible.

## 4 Methods

### 4.1 Study design and data-use declaration

This was a secondary analysis of de-identified controlled-access data from PPMI and PDBP. The PPMI protocol was approved by the institutional review board or independent ethics committee at each participating site, and all participants provided written informed consent [2, 3]. PDBP contributing studies obtained local institutional review board or ethics approval and written informed consent under their parent protocols [21]. No new participant was recruited and no new biospecimen was collected for this analysis. PPMI analyses used the 20 December 2025 data release; local PDBP exports were analyzed under the PDBP data-use agreement. Dataset provenance was recorded before analysis, and participant-level source data are not redistributed.

The prespecified progression analysis defined the 12-month endpoint, participant-level split unit, ridge comparator, source-age handling, decision thresholds and excluded claims. Enrollment-era, strict-calendar and site-disjoint evaluations were conducted without tuning the model family, regularization or feature blocks to their outcomes. The selective-UQ and semisynthetic protocols were finalized on 17 August 2026. Before outcome-facing runs, the finalized implementation required mutually exclusive participant roles, participant-level conformal scores and leakage checks based on observed participant overlap; only results from these specifications are reported.

The analysis sequence was: (i) the rolling PPMI source-time benchmark; (ii) selective uncertainty on the same endpoint with independent fitting and calibration roles; (iii) a semisynthetic audit of the decision rule; and (iv) PDBP clinical transport and imaging-feasibility audits. Populations and endpoints were not pooled, and secondary stress tests could not override the primary result.

### 4.2 PPMI dated spine and quality control

Participant Status defined the PD population. MDS-UPDRS Part III examination or information dates were resolved to calendar month. Duplicate participant-month rows were collapsed by median for numeric measures, with source-row counts retained for audit. DaT-SPECT, MoCA, medication intervals, demographics and diagnosis dates were linked by participant and observed month. Disease duration was the non-negative interval from diagnosis month to cutoff month.

This monthly collapse can combine examination states. Side sums and laterality were calculated per source row before their monthly medians were taken; a median of ratios need not equal a ratio of median sums. The primary benchmark is unchanged by the descriptive case audit. In P1-B, the two ratios 1.00 (OFF) and 0.75 (ON) give the saved monthly value 0.875. State-restricted sensitivities, rather than relabelling that median as one examination, address the resulting ambiguity.

For regional DaT-SPECT, left and right SBRs were converted to numeric values. Derived mean and asymmetry were retained only when both regional SBRs were non-negative and their sum was positive. This requirement keeps the normalized asymmetry within its physically interpretable range. An invalid asymmetry was represented as missing rather than clipped. No observation after a cutoff was used to impute, scale, select or construct a predictor for that cutoff.

### 4.3 Rolling-origin endpoint

For every eligible clinical cutoff, the endpoint was the MDS-UPDRS Part III [22] assessment closest to 12 months within a plus or minus 3-month tolerance. The change score was annualized by the observed month interval. Eligibility required at least three unique clinical months and at least 12 months of prior clinical history. Clinical and MoCA slopes required at least three observations over at least 12 months. DaT-SPECT slope allowed two observations over at least 12 months because imaging was sampled less frequently.

The primary endpoint was annualized 12-month MDS-UPDRS Part III change. This choice forecasts worsening relative to the participant’s current motor state rather than reconstructing a future absolute score that is dominated by the current score. Twenty-four-month and thresholded-worsening screens were treated as exploratory because they did not improve the transport conclusion.

### 4.4 Information blocks and estimators

The algorithm has three separate outputs. First, dated records form a cutoff-specific feature vector, and a participant-held-out ridge pipeline forecasts annualized motor change. Second, the same pre-cutoff record can be compressed into interpretable evidence coordinates; those are descriptions of available support, not additional independent measurements. Third, a separately fitted uncertainty model estimates residual risk and is calibrated on disjoint participants. Figure 1a–c shows the record-to-representation steps; panel d distinguishes the primary forecasts from the uncertainty model’s own mean and interval. Figure 3b–e then tests added value, coverage, selective discrimination and transport separately. Failure at one stage cannot be repaired by renaming a representation or displaying a favorable patient.

The clinical trajectory and context block comprised current MDS-UPDRS Part III, signed laterality, gait, freezing, active LEDD, number and span of prior visits, motor and laterality slopes, age, recorded sex code and diagnosis-derived disease duration. The dated source-value block comprised putamen mean SBR, signed putamen asymmetry, putamen SBR slope, MoCA [23] and MoCA slope. The observation-process block comprised DaT-SPECT and MoCA ages plus indicators for availability of each level and slope. The full source-time model combined all blocks.

This source organization is part of the reusable data-science contribution: each value retains a participant, biological or clinical role, observation time and availability status before it becomes a predictor or evidence summary. The same dated records support different questions without treating every collected modality as a fitted input. Genomic, transcriptomic and direct neural-circuit dynamics are not estimated in the present primary benchmark.

Figure 1 reuses saved observations, source ages, evidence scores, reliability weights and held-out predictions without refitting or reselection. Missing sources occupy a separate strip rather than being plotted as zero-valued measurements.

The reference human and brain in Figure 2 locate the measured body side and putamen; neither surface is reconstructed from a participant. Supplementary Section S12 separates selected reference motor, associative and nigrostriatal routes from the non-localizing cognitive and fluid sources. Human connectivity studies inform this anatomical context [18]; connectivity itself is not measured in these cases.

Each candidate used the same pipeline: training-partition median imputation with missingness indicators, robust scaling and ridge regression with alpha=10. No feature selection used test outcomes. The prespecified internal split assigned participants to five folds after stratification by participant median outcome; all rows from one person remained together. The non-stratified GroupKFold sensitivity was generated independently.

Outcome-blind sensitivity analyses either assigned each participant equal total fitting weight or retained only the earliest or latest eligible cutoff. Medication-state analyses added cutoff ON/OFF indicators to both candidate and reference models and separately summarized ON-to-ON, OFF-to-OFF, switched-state and LEDD-observed subsets. Outcome examination state defined sensitivity strata only and was never used as a predictor.

### 4.5 Enrollment-era, calendar-deployment and site-disjoint evaluations

For each eligible participant, the earliest dated PD motor observation in the full clinical spine was selected. Dates through 31 December 2019 defined the development era; dates from 1 January 2020 defined the temporal test era. Every model, imputer and scaler was fit only in the development era and applied once to the later era. No later-era participant was used for feature transformation, model selection, refitting or recalibration. This is a within-program temporal and protocol stress test, not external validation.

For the stricter calendar-deployment sensitivity, development rows were further restricted to outcomes completed before 1 January 2020. Thus neither the predictor cutoff nor its 12-month endpoint crossed the deployment boundary. The untouched test remained the 442 participants first observed from 2020.

Site identifiers were obtained from the PPMI curated data cut dated 12 June 2023 and linked by participant identifier. The 951 overlapping benchmark participants represented 52 sites. Sites were sorted by participant count and greedily assigned to the currently smallest of five folds; outcomes were not used for assignment. Every cutoff from a participant and every participant from a site remained in one test fold. As a descriptive shift audit, one earliest cutoff per participant was used in five-fold logistic regression to classify enrollment era from predictor history. Its AUC interval used 10,000 participant bootstrap replicates. The classifier quantified cohort distinguishability and was not interpreted as a clinical model.

### 4.6 Performance and participant-cluster uncertainty

We reported *R*^2^, MAE, RMSE, Spearman correlation, and calibration intercept and slope. Candidate comparisons were paired on identical cutoff rows. Confidence intervals used 10,000 participant-cluster bootstrap replicates: participants were sampled with replacement and every cutoff belonging to a sampled person was retained. Positive Δ*R*^2^ and negative delta MAE favor the candidate. The prespecified added-value criterion required Δ*R*^2^ of at least 0.02, a 95% interval above zero and delta MAE below zero.

For endpoint-noise sensitivity, median absolute error was calculated from the unchanged held-out predictions. In a second analysis, the motor-change outcome was bounded at the 1st and 99th percentiles estimated only in each training partition. Imputation, scaling and ridge regression were then refitted to the bounded training outcome; held-out outcomes and predictions were clipped using those training limits. Internal folds and the earlier-to-later calendar split were otherwise unchanged. Five thousand participant-cluster bootstrap draws compared the full and clinical models. These analyses could preserve or weaken the verdict but could not replace the primary endpoint or decision threshold.

### 4.7 Source-aware selective uncertainty

The selective-UQ protocol was finalized on 17 August 2026 before model fitting. The predictive mean retained the full source-time ridge pipeline. Because participants contributed different numbers of rolling cutoffs, mean and uncertainty estimators used inverse-cutoff weights so each participant had equal total fitting weight. Mean prediction was promoted only when the added-value criterion passed without an unfavorable site or calendar reversal; the observed internal magnitude and calendar interval did not satisfy that rule.

Every source retained value, observation time, age in months and availability. DaT-SPECT reliability was availability multiplied by exp(*−*age*/*24); MoCA reliability used exp(*−*age*/*12). Ages were floored at zero; missing ages were assigned 120 months before multiplication by availability. With *c*(*x*) = min(1, max(0*, x*)), clinical-history reliability was

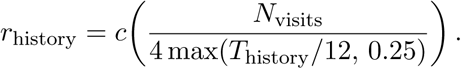

Fixed-scale levels were *m* = *c*(UPDRSIII*/*132), *d* = *c*((3 *−* SBR)*/*3), and *q* = *c*((30 *−* MoCA)*/*30). Disagreement inputs were *|m − d|*, *|m − q|*, and

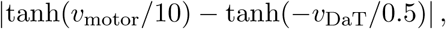

where velocities were per year. The model also received the mean of the three reliability variables and the mean of the available disagreement variables. Missing disagreement values remained missing before training-partition imputation. These fixed-scale formulas are reliability and disagreement proxies, not calibrated biological probabilities.

Every outer training set was partitioned by participant into three mutually exclusive development roles. A deterministic 20% subset, with at least 100 participants, supplied conformal calibration. One quarter of the remaining participants, with at least 50, fitted the residual-scale model. All other development participants fitted the predictive mean. The outer test participants occupied none of these roles. Participant-disjoint folds contained 507 mean-fit, 169–170 scale-fit, 170 calibration and 211–212 test participants per fold; site-disjoint folds contained 456, 152, 152–153 and 190–191, respectively; the strict-calendar analysis contained 348, 116, 116 and 442. The same mean model generated predictions for scale fitting, calibration and testing, so neither residual-scale fitting nor calibration reused a participant that fitted the mean. Inverse-cutoff weights gave every participant equal total weight within the mean-fit and scale-fit roles.

Clinical uncertainty used clinical trajectory, context and the predicted mean. Generic uncertainty added raw source values, ages and availability. Source-aware uncertainty further added reliability and disagreement coordinates. All three used the same histogram gradient-boosting residual model with learning rate 0.05, 160 iterations, seven terminal leaves, minimum leaf size 40 and L2 penalty 2. The architecture and seed were fixed before held-out risk–coverage results.

For each calibration participant, we divided every absolute residual by its predicted scale and retained that participant’s maximum normalized residual. The finite-sample higher quantile of these participant maxima formed the 90% interval multiplier. This cluster split-conformal construction targets simultaneous coverage over all eligible cutoffs from an exchangeable participant, not merely average row coverage. We therefore reported both row coverage and the proportion of participants for whom every interval covered, along with width, source-availability and severity strata, and uncertainty–error Spearman correlation.

Selective risk was MAE after ranking predictions from lowest to highest uncertainty. The primary area under the risk–coverage curve (AURC) integrated risk over 50%–100% retention and divided by the interval length. Five thousand participant-bootstrap draws compared source-aware with clinical AURC. Promotion required an upper 95% bound below zero, no unfavorable strict-calendar reversal, row and simultaneous participant coverage of at least 0.87 overall, the same row-coverage floor in every source-availability stratum with at least 50 participants, and no participant overlap among fitting, calibration and test roles. Site-disjoint folds retained entire sites, and strict-calendar training admitted only earlier-participant rows whose outcomes occurred before 1 January 2020. Failure of any requirement returned `ABSTAIN`, a research-validity status rather than a clinical action.

The semisynthetic audit generated 2,000 replicates for each of four scenarios spanning null effects, stable effects, repeated-participant leakage and calendar reversal. Each replicate sampled 200, 400 or 800 participants per cohort; two, three or four visits per participant (at least six under leakage); participant-effect SD 0.15, 0.40 or 0.70; source missingness 0.05, 0.25 or 0.50; and stable source-risk coefficient 0.30, 0.55 or 0.80. Calendar reversal used the negative full effect, negative half effect or zero, and each internal interval used 200 participant-cluster bootstrap draws. Four independently generated cohorts supplied residual-scale fitting, conformal calibration, internal testing and calendar testing. The leakage scenario deliberately reused identities across roles and added a learned participant-specific residual. Scenario labels were hidden from the decision rule, which could use only the internal AURC interval, row and participant coverage, calendar direction and directly measured participant intersections. The naive rule required internal improvement and adequate coverage; the complete gate additionally prohibited overlap and calendar reversal. These simulations audit gate behavior, not PD biology.

### 4.8 Cohort representation and descriptive subgroup audit

Baseline characteristics used each participant’s earliest eligible rolling cutoff. Age, recorded sex, diagnosis-derived disease duration, current motor total and active LEDD came from the dated spine; PPMI race and ethnicity came from the curated demographics module. Race fields were reported as recorded and were not forced into mutually exclusive categories.

The subgroup audit reused the primary out-of-fold predictions and did not refit or select models within a stratum. Age at the earliest cutoff was grouped as below 60, 60-69 or 70 years and older; recorded sex was summarized as female or male according to the source coding. Paired Δ*R*^2^ and delta MAE intervals used 10,000 participant-cluster bootstrap draws within each subgroup. These secondary analyses were descriptive; no interaction hypothesis, fairness threshold or subgroup-specific clinical claim was specified.

### 4.9 Continuous evidence composition

Evidence coordinates were calculated independently inside each outer fold. For feature *j*, the *n_j_* finite, available training values defined a smoothed midrank percentile,

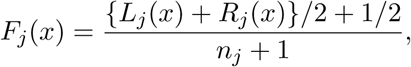

where *L_j_*(*x*) and *R_j_*(*x*) count training values below *x* and at or below *x*, respectively. A feature remained missing if the input was unavailable or fewer than ten reference values existed. The oriented evidence was *e_j_* = *d_j_*[2*F_j_*(*x_j_*) *−* 1], with fixed direction *d_j_*. Observation age in months was clipped to [0,48] and weighted by *w_j_* = exp[*−* log(2)age*_j_/*24], a 24-month half-life. A coordinate was the weighted mean over available features with finite values and ages; no valid feature yielded a missing coordinate. Its separate reliability proxy was the sum of valid weights capped at one. Thus source age changes relative weights within a multi-source coordinate, but cancels from a single-source normalized mean; reliability and feature count must be inspected separately. These evidence weights differ from the exponential reliability variables used by the selective-UQ model.

The dopaminergic coordinate used lower putamen SBR; injury/glial used higher NfL and GFAP; co-pathology/cognitive used lower amyloid-beta42, higher p-tau, t-tau and p-tau217, and lower MoCA; lysosomal used lower GCase activity. The continuous model added four evidence coordinates, four reliability coordinates and all six pairwise coordinate products to the clinical reference inputs. Products requiring a missing coordinate remained missing before training-partition imputation. The hard-state comparator instead added four one-hot indicators for the largest available coordinate; all four were zero if none was available. Bulk CSF alpha-synuclein was excluded as an aggregation anchor. Scores were relative evidence summaries and were never interpreted as calibrated probabilities.

### 4.10 Outcome-blind illustrative records

The patient-display analysis was specified on 11 September 2026 after the primary analyses and is descriptive. Each participant contributed their earliest eligible rolling cutoff, with at least two scored biological coordinates. Candidate pairs shared recorded sex and outer fold and differed by no more than three motor points, five years of age, two years of disease duration and two annualized points of prior motor slope. One record had DaT-SPECT and MoCA ages no greater than twelve months; the other had absent DaT-SPECT, DaT-SPECT older than twenty-four months, absent MoCA or MoCA older than twelve months. These are display strata, not clinical freshness thresholds. The minimum sum of squared caliper-scaled clinical differences selected one pair; a deterministic hash resolved ties. Thirty pairs met these rules. Future values, forecast errors, interval coverage and prediction changes were excluded from selection. Saved source measurements, training-fold evidence scores and model-specific held-out predictions were then joined by participant and cutoff; source details are reported in Supplementary Section S11. All observed trajectories use actual relative months. Pair similarity is not causal matching, and different treatment documentation remains visible rather than being interpreted as a treatment contrast.

### 4.11 Learned-model negative controls

Reconstruction error was expressed as standardized mean absolute error (sMAE). Each held-out absolute error was divided by that target’s sample standard deviation in the corresponding baseline-model fitting partition, excluding calibration and test participants. The same fold-local reference scale was applied to the learned model. Errors were averaged within target and then equally across targets in each masked source; the display reports point summaries, not confidence intervals.

Whole-source masking removed target-source values, availability indicators and observation-age features together. Transparent and learned candidates used the same participant folds, and conformal intervals used disjoint calibration participants. The partial-view model projected each dated feature-and-observation channel to 64 dimensions, encoded the 96-month panel with a one-layer 64-unit GRU, and combined the final hidden state, latest snapshot and demographics before estimating five non-exclusive evidence coordinates and a 24-dimensional nuisance state. A sign-constrained linear decoder enforced prespecified anchor directions; unconstrained cross-loadings and nuisance loadings were penalized. Training used AdamW with learning rate 5 *×* 10*^−^*^4^, weight decay 10*^−^*^4^, batch size 192, dropout 0.15, at most 180 epochs and participant-disjoint early stopping with patience 25. Whole-source ablations removed DaT-SPECT, molecular, digital or provenance blocks and were compared on identical held-out participants.

As a secondary temporal stress test, the contextual temporal candidate used a 96-month masked panel, patch lengths 8, 16 and 32, width 64, depth two, dropout 0.10 and a fold-normalized static-context residual. AdamW used learning rate 3 *×* 10*^−^*^4^, weight decay 10*^−^*^4^, batch size 128 and at most 175 epochs with patience 25. Fifty fixed initializations were trained on the 616 earlier-era participants; early stopping used a participant-disjoint validation subset drawn only from that era. Each model was evaluated once in the 442 later-era participants. All seeds, ranges and comparisons were retained; no seed was selected and no ensemble was formed.

### 4.12 PDBP clinical transport

PDBP clinical transport was restricted to participants whose final available diagnosis was exactly Parkinson’s disease. Timing used supplied elapsed days; calendar dates were not inferred. Harmonized predictors were motor total, signed laterality, absolute laterality, gait, freezing and global bradykinesia. The model was developed in PPMI, then linearly recalibrated using 77 prespecified PDBP calibration participants and evaluated once in 156 untouched participants. The external criterion required calibration slope 0.7-1.3 and an upper 95% confidence bound no more than 0.10 MAE worse than a calibration-mean reference.

### 4.13 PDBP image and diagnosis audit

The complete 3.49-GiB local PDBP SPECT bundle was audited by indexing the central directory of the outer archive without opening or processing participant images. Catalog entries were joined by participant and visit to archive members, DaTSCAN modality labels, raw and curated timed MDS-UPDRS records and exact neurological-examination diagnoses. Eligibility required a final diagnosis exactly equal to Parkinson’s disease, a present image reference and a timed clinical record in the same participant. The audit characterizes availability and domain overlap only; no PDBP SBR or image-derived feature was estimated.

### 4.14 Statistics and reproducibility

All tests were two-sided and confidence intervals were 95% intervals. Repeated cutoffs were handled with participant-level splitting and clustered resampling. The primary random seed was 20260725; temporal neural initializations used 20260803–20260852, and selective-UQ and semisynthetic analyses used seed 20260817. Selective-UQ AURC intervals used 5,000 participant-bootstrap draws; simulations used 2,000 replicates per scenario and 200 participant-cluster bootstrap draws per replicate.

Analyses used Python 3.11, pandas, NumPy, SciPy, scikit-learn, matplotlib and PyTorch. Model families and complete seed sets were specified independently of held-out outcomes. Programmatic checks verified source chronology, participant and site separation, disjoint uncertainty roles, conformal scoring, calendar boundaries, asymmetry bounds and PDBP cohort eligibility. Reporting was informed by TRIPOD+AI and STROBE principles [12, 24]. The Supplementary Information reports supporting populations, sensitivity analyses, model-capacity controls, uncertainty results, simulations and transport audits; all analytical methods are specified above.

### 4.15 Use of AI-assisted tools

AI-assisted tools, including OpenAI Codex, were used only for manuscript revision, code development and debugging. The authors reviewed and verified all outputs and retain full responsibility for the manuscript, analyses and code.

## 5 Extended Patient Walkthroughs

### 5.1 What is visible before a forecast is made?

The two patients in Figures 1 and 2 illustrate a practical problem in reviewing multimodal models: similar motor summaries do not imply equivalent evidence. The first question is not whether one displayed prediction is closer to its outcome. It is whether the record retains enough information to tell what was measured, when it was measured and what interpretation that measurement can support.

For P1-A, the cutoff motor total is 10 and the prior slope is 1.29 points/year, based on three clinical months spanning 17 months. Its single cutoff examination contains four left-side paired motor points and no right-side paired points. The remaining motor-total points arise from items outside the paired laterality sum. The scan in that month has right/left putamen binding 0.71/0.96. This provides two compatible but distinct observations: a left-predominant examination and relatively lower right putamen binding. Neither observation proves that every symptom is dopaminergic or that the apparent imbalance measures global disease severity.

For P1-B, the cutoff total is 11 and the prior slope is 1.50 points/year, based on four clinical months spanning 16 months. There are two cutoff examinations. The OFF record has left/right paired sums 0/8; the ON record has 1/7. Both totals are 11, but the distributions are not identical. The frozen monthly spine uses numeric medians, giving laterality 0.875. It does not select the more favorable examination, and it does not interpret the difference as a medication effect. No observed DaT-SPECT measurement is available in this rolling record, so a right-predominant examination cannot be relabelled as an observed left-brain deficit.

This distinction between a single source observation and an aggregate is important even when the final number is unchanged. An apparent difference between patients could reflect phenotype, acquisition practice, treatment context, or several factors together. The display makes these alternatives inspectable without claiming to identify their cause.

### 5.2 From measured values to evidence and reliability

For an observed source value *x_j_*, the evidence transformation uses only its training-fold empirical distribution:

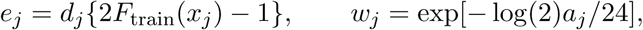

where *d_j_* fixes the biological direction and *a_j_* is the source age in months, capped at 48 in the source panel. For available sources in coordinate *k*,

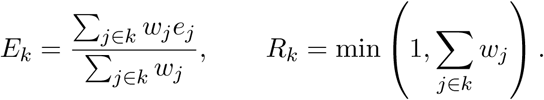

An empty source set has no evidence coordinate; it is not assigned a normal value. *E_k_* summarizes relative evidence and *R_k_* is a separate availability/age proxy. Neither is a calibrated probability or diagnostic severity scale. In particular, a negative dopaminergic score means weaker relative evidence within the training distribution, not normal dopamine-transporter binding.

P1-A has a dopaminergic coordinate of approximately -0.564 supported by contemporaneous imaging, and a cognitive coordinate of 0.942 supported by MoCA 19. Its injury/glial and lysosomal coordinates are missing. P1-B has cognitive evidence 0.507 from MoCA 25, injury/glial evidence 0.343 supported by NfL, and lysosomal evidence 0.540 supported by GCase; its dopaminergic coordinate is missing. These names identify the interpretation attached to measured proxies. NfL does not diagnose inflammation, MoCA does not establish a molecular co-pathology, and GCase activity does not establish a GBA mutation.

The older GCase measurement provides a useful worked check. At 14 months, its weight is 2*^−^*^14^*^/^*^24^ *≃* 0.667. Because it is the only available source for that coordinate, the weight cancels from the normalized mean: *E_k_* = *w_j_e_j_/w_j_* = *e_j_*. Its score therefore remains 0.540 while its reliability falls to 0.667. Lowering the score solely because it is old would be a different algorithm. Displaying score and reliability separately preserves the distinction between what was measured and how current that support is. The selective-uncertainty model uses different exponential time constants, specified in Methods; these two weighting systems are not interchangeable.

### 5.3 Regional context without invented localization

Figure 7a locates selected routes relevant to interpreting striatal measurements, informed by human connectivity research [18]. Panel b has a different evidential status: its source dates and missing values are actual patient observations. The model does not measure cortical connectivity, localize a MoCA result to the middle frontal gyrus, or infer a nigral lesion from NfL. The regional diagram explains why an imaging observation is anatomically meaningful without promoting a non-localizing biomarker into a patient-specific brain map.

**Figure 7:**
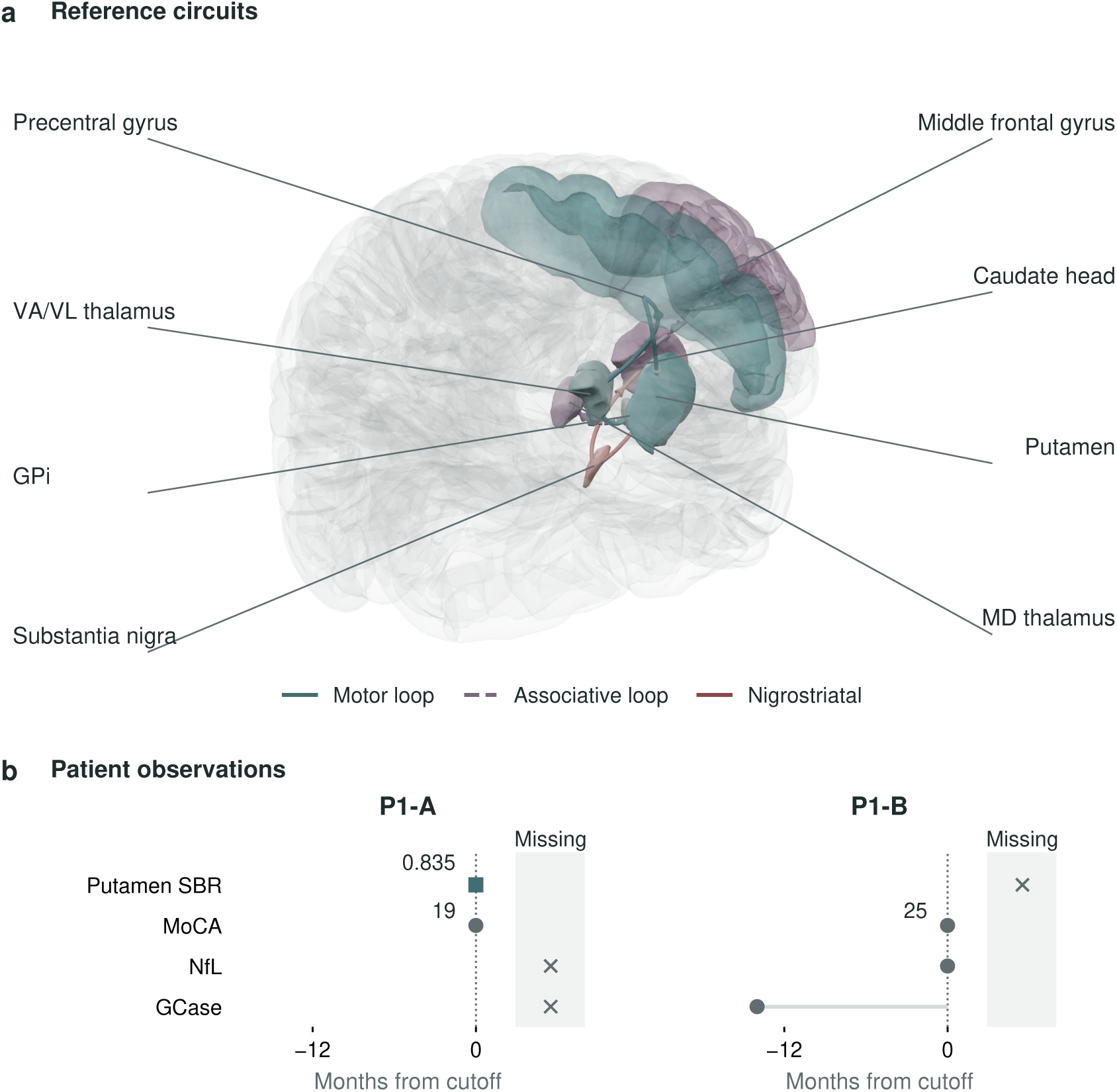
An anatomical map is context, not an additional patient measurement. a, Selected left-hemisphere motor (solid teal), associative (dashed mauve) and nigrostriatal (rust) routes on named reference structures. GPi is internal globus pallidus; VA/VL/MD are ventral anterior, ventral lateral and mediodorsal thalamus. Routes are schematic and incomplete; they do not show patient connectivity, excitation, inhibition or disease propagation. b, Actual dated source support for P1-A/P1-B, including absent sources. Putamen SBR is a bilateral mean here. The left-hemisphere reference view must not be mistaken for either patient’s affected hemisphere; MoCA, NfL and GCase do not localize dysfunction to the highlighted structures. Atlas: NIH 3D 3DPX-020960, Allen Human Reference Atlas, CC BY 4.0.

### 5.4 From a current record to an honest future comparison

For each cutoff *t*, only observations dated at or before *t* enter the predictors. The selected future motor summary *Y_t_*_+_*_h_*, with 9 *≤ h ≤* 15 months, defines

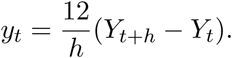

For P1-A, 12(22 *−* 10)*/*9 = 16 annualized points/year; for P1-B, 12(19 *−* 11)*/*12 = 8. Annualization does not relocate the nine-month examination to month twelve and does not imply linear biological change between visits. Prior slopes are predictors; these later outcomes are not.

The two primary clinical forecasts are 4.447 and 1.759 points/year; the full source-time forecasts are 7.317 and 2.463. Adding sources moves both forecasts toward the eventual observed change in these examples, but that outcome was not used in their selection. The separate nested uncertainty fit has means 9.064 and 3.006, with approximately 90% intervals [0.020,18.109] and [-17.547,23.558]. Those intervals belong to the uncertainty model, not to the primary ridge or continuous-evidence forecasts. Their width is a substantive result: the representation is interpretable, but these individual trajectories are not precisely resolved.

The two examples demonstrate traceability, not the average value of imaging. Cohort-level comparisons supply that answer. The full source-time increment is 0.015 in internal *R*^2^, below the 0.02 magnitude criterion, and its strict-calendar interval includes zero. A favorable movement in a selected forecast cannot overturn either result.

### 5.5 What a passed gate would permit

Figure 3 separates four questions that are often conflated. An internal paired gain asks whether dated sources contain information beyond the reference in that evaluation. Calendar testing asks whether that gain survives a later acquisition era without refitting. Simultaneous participant coverage asks whether all evaluated cutoffs for a person are contained in their intervals. Selective prediction asks whether uncertainty can identify a lower-error subset. The external panel tests a separately harmonized clinical model, not the full multimodal representation.

These distinctions change how the result should be used. A covered but very wide interval is not precise prognosis. A small positive internal increment is not temporal transport. A score named for a mechanism is not causal identification. The useful output for a research team is an inspectable evidence state accompanied by the particular claims that passed, failed or remain untested. In this study that record retains the real-data failures, the residual failures in semisynthetic stress tests, and the absence of treatment-policy evidence. It is a foundation for a better specified next experiment, not a substitute for one.

## Data Availability

All data produced in the present study are available upon reasonable request to the authors

https://cvc-lab.github.io/projects/pd-research-companion/pd-validity-gates/

## 6 Data availability

PPMI data were obtained under the PPMI data-use agreement from www.ppmi-info.org/access-data-specimens/download-data (RRID:SCR_006431). PDBP data were obtained under the NINDS PDBP data-use agreement from pdbp.ninds.nih.gov. These participant-level resources cannot be redistributed by the authors. Authorized researchers may request access directly through the program portals. Release-specific module manifests, variable crosswalks, code and non-identifying aggregate metrics permit authorized users to reconstruct the analyses. No new primary human-participant data were generated.

## 7 Code availability

Analysis code and supporting documentation are maintained at https://github.com/CVC-Lab/pd-validity-gates. For repository access, please contact the authors. Controlled participant-level data are not redistributed and must be obtained directly from PPMI and PDBP under their respective data-use agreements.

## 8 Acknowledgements

This research was supported in part by grants and gifts from the Peter O’Donnell Foundation, the Michael J Fox Foundation, Jim Holland-Backcountry and Michael-Connie Rasor Foundations towards curing Parkinson’s Disease.

Data used in the preparation of this article were obtained on 2025-12-20 from the Parkinson’s Progression Markers Initiative (PPMI) database (www.ppmi-info.org/access-data-specimens/download-data), RRID:SCR_006431. For up-to-date information on the study, visit www.ppmi-info.org. PPMI is a public-private partnership funded by The Michael J. Fox Foundation for Parkinson’s Research and funding partners, including 4D Pharma, AbbVie, AcureX, Allergan, Amathus Therapeutics, Aligning Science Across Parkinson’s, AskBio, Avid Radiopharmaceuticals, BIAL, BioArctic, Biogen, Biohaven, BioLegend, BlueRock Therapeutics, Bristol Myers Squibb, Calico Labs, Capsida Biotherapeutics, Celgene, Cerevel Therapeutics, Coave Therapeutics, DaCapo Brainscience, Denali, Edmond J. Safra Foundation, Eli Lilly, Gain Therapeutics, GE HealthCare, Genentech, GSK, Golub Capital, Handl Therapeutics, insitro, Jazz Pharmaceuticals, Johnson & Johnson Innovative Medicine, Lundbeck, Merck, Meso Scale Discovery, Mission Therapeutics, Neurocrine Biosciences, Neuron23, Neuropore, Pfizer, Piramal, Prevail Therapeutics, Roche, Sanofi, Servier, Sun Pharma Advanced Research Company, Takeda, Teva, UCB, Vanqua Bio, Verily, Voyager Therapeutics, the Weston Family Foundation and Yumanity Therapeutics.

Data and biospecimens used in preparation of this manuscript were obtained from the Parkinson’s Disease Biomarkers Program (PDBP) Consortium, supported by the National Institute of Neurological Disorders and Stroke at the National Institutes of Health. Investigators include Roger Albin, Roy Alcalay, Alberto Ascherio, Thomas Beach, Sarah Berman, Bradley Boeve, F. DuBois Bowman, Shu Chen, Alice Chen-Plotkin, William Dauer, Ted Dawson, Paula Desplats, Richard Dewey, Ray Dorsey, Jori Fleisher, Kirk Frey, Douglas Galasko, James Galvin, Dwight German, Steven Gunzler, Lawrence Honig, Xuemei Huang, David Irwin, Un Kang, Kejal Kantarci, Anumantha Kanthasamy, Daniel Kaufer, Horacio Kaufmann, Qingzhong Kong, James Leverenz, Allan Levey, Carol Lippa, Irene Litvan, Oscar Lopez, Jian Ma, Richard Mailman, Lara Mangravite, Karen Marder, Kelly Mills, Nandakumar Narayanan, Laurie Orzelius, Vladislav Petyuk, Judith Potashkin, Liana Rosenthal, Rachel Saunders-Pullman, Clemens Scherzer, Michael Schwarzschild, Nicholas Seyfried, Tanya Simuni, Andrew Singleton, David Standaert, Debby Tsuang, David Vaillancourt, Jerrold Vitek, David Walt, Andrew West, Cyrus Zabetian and Jing Zhang. The PDBP Investigators have not participated in reviewing the data analysis or content of this manuscript.

We thank the participants and investigators of PPMI and PDBP. Computational analyses used resources of the Texas Advanced Computing Center at The University of Texas at Austin.

## 9 Author contributions

H.T. and P.Y. contributed equally. H.T.: conceptualization, methodology, software, formal analysis, validation, visualization, writing: original draft, and writing: review and editing. P.Y.: conceptualization, data curation, methodology, software, formal analysis, validation, visualization, writing: original draft, and writing: review and editing. C.B.: conceptualization, methodology, supervision, project administration, resources, and writing: review and editing. All authors reviewed and approved the manuscript.

## Supplementary Information

### Supplementary Results

#### S1. Analysis populations

| Analysis layer | Rows or cutoffs | Independent participants | Purpose |
| --- | --- | --- | --- |
| Dated PPMI clinical spine | 25,118 patient-months | 4,773 all statuses; 1,602 PD | Date and source audit |
| PPMI baseline 12-month resource | 1,256 outcomes | 1,256 PD | PDBP-harmonized internal reference |
| PPMI slope-eligible resource | Not additive | 1,348 PD | At least 3 motor visits over at least 12 months |
| PPMI rolling-origin benchmark | 5,404 cutoffs | 1,058 PD | Primary future motor-change analysis |
| Development era through 2019 | 4,664 cutoffs | 616 PD | Temporal model development |
| Post-2020 era | 740 cutoffs | 442 PD | Untouched temporal test |
| Strict pre-2020 outcome training | 3,575 cutoffs | 580 PD | Strict calendar sensitivity |
| Site-disjoint overlap | 5,281 cutoffs | 951 PD across 52 sites | Acquisition-site challenge |
| PDBP confirmed-PD outcomes | 233 outcomes | 233 PD | External clinical transport |
| PDBP calibration / untouched test | 77 / 156 | 77 / 156 PD | Recalibration and final evaluation |
| PDBP DaTSCAN catalog | 1,045 records | 139; none eligible idiopathic PD | Imaging-domain feasibility audit |

These rows overlap within PPMI and are not additive. The effective generalization sample for the rolling benchmark is 1,058 people, not 5,404 independent observations.

#### S2. Participant characteristics and representativeness

Characteristics use one earliest eligible cutoff per participant. Active LEDD percentages use only participants with recorded exposure; unavailable exposure was never coded as untreated.

| Cohort | Participants | Age, mean (SD), y | Female, n (%) | Disease duration, median (IQR), y | Motor total, mean (SD) | Active LEDD / recorded, n/N (%) |
| --- | --- | --- | --- | --- | --- | --- |
| PPMI rolling benchmark | 1,058 | 63.8 (9.7) | 407 (38.5%) | 1.8 (1.4-2.8) | 23.5 (10.9) | 775/792 (97.9%) |
| First observed through 2019 | 616 | 62.9 (9.9) | 249 (40.4%) | 1.8 (1.3-3.1) | 23.2 (11.2) | 479/488 (98.2%) |
| First observed 2020 onward | 442 | 65.2 (9.2) | 158 (35.7%) | 1.8 (1.4-2.5) | 24.0 (10.5) | 296/304 (97.4%) |

Age was available for 1,055 of 1,058 participants. Source labels follow the data dictionaries and do not imply measured gender identity. PPMI permitted multiple race selections: 1,011 participants (95.6%) were recorded White, 20 (1.9%) Black or African American, 17 (1.6%) Asian, 5 (0.5%) American Indian or Alaska Native, 1 (0.1%) Native Hawaiian or other Pacific Islander, 22 (2.1%) other race and 6 (0.6%) unknown; 53 (5.0%) were recorded Hispanic or Latino. Sparse non-White cells did not support stable race-stratified evaluation, so no demographic-fairness claim is made.

#### S3. Source chronology and observation process

| Enrollment era | People | Cutoffs | Motor change, mean (SD) | DaT level | DaT slope | MoCA level | MoCA slope | Active LEDD |
| --- | --- | --- | --- | --- | --- | --- | --- | --- |
| First observed through 2019 | 616 | 4,664 | 1.31 (9.39) | 97.0% | 81.3% | 99.8% | 75.3% | 93.3% |
| First observed 2020 onward | 442 | 740 | 0.34 (8.95) | 98.1% | 80.4% | 99.9% | 42.0% | 76.5% |

The era definition is an operational proxy based on the earliest dated PD motor observation. It is not an adjudicated recruitment date. A cross-validated logistic model using one earliest predictor history per participant distinguished the eras with AUC 0.978 (95% participant-bootstrap CI 0.969-0.987). The largest standardized shift was prior visit count (standardized mean difference -1.91). This is a dataset-shift diagnostic, not a disease classifier.

#### S4. Primary future-only benchmark

| Model | $R^2$ (95% CI) | MAE (95% CI) | RMSE (95% CI) | Spearman rho | Calibration slope |
| --- | --- | --- | --- | --- | --- |
| Clinical trajectory and context | 0.102 (0.081, 0.121) | 6.766 (6.585, 6.957) | 8.846 (8.576, 9.124) | 0.301 | 0.958 |
| Clinical plus dated source values | 0.117 (0.096, 0.136) | 6.706 (6.528, 6.895) | 8.770 (8.511, 9.045) | 0.321 | 0.939 |
| Clinical plus observation process | 0.102 (0.082, 0.121) | 6.761 (6.581, 6.951) | 8.845 (8.579, 9.121) | 0.301 | 0.950 |
| Full source-time history | 0.118 (0.097, 0.136) | 6.706 (6.528, 6.896) | 8.769 (8.511, 9.044) | 0.320 | 0.936 |

| Candidate versus clinical reference | $\Delta R^2$ (95% CI) | Delta MAE (95% CI) | Gate interpretation |
| --- | --- | --- | --- |
| Dated source values | 0.015 (0.006, 0.025) | -0.061 (-0.100, -0.021) | Interval-supported but below 0.02 threshold |
| Observation process | 0.000 (-0.003, 0.003) | -0.005 (-0.017, 0.007) | No internal increment |
| Full source-time history | 0.015 (0.006, 0.025) | -0.060 (-0.101, -0.020) | Mean-prediction gate failed |

The non-stratified GroupKFold sensitivity gave Δ*R*^2^ 0.018 (95% CI 0.008-0.028) and delta MAE -0.068 (-0.108 to -0.025). The conclusion did not depend on outcome-stratified fold balancing.

#### S5. Repeated-cutoff, site and calendar challenges

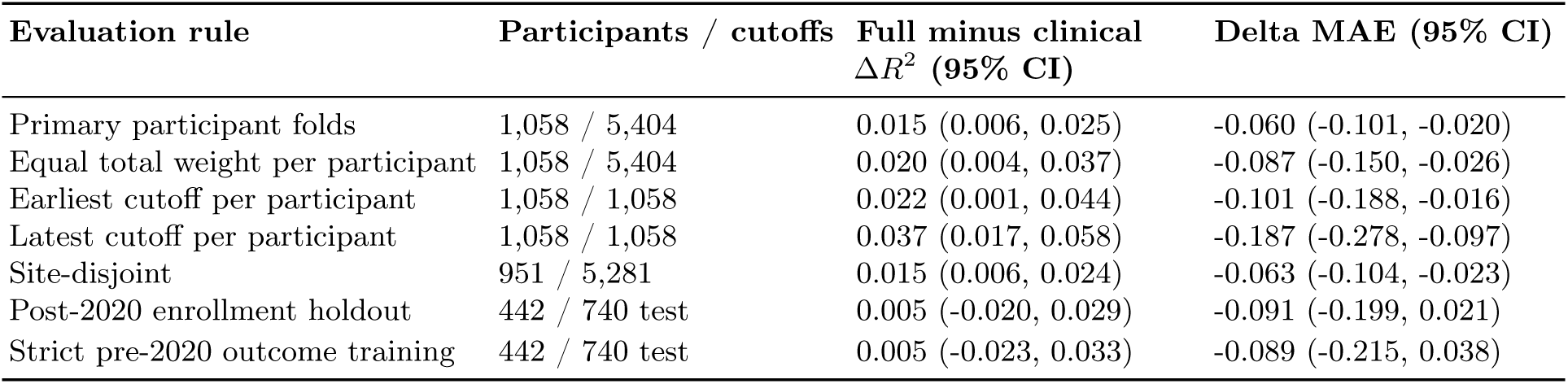

Under the strict calendar design, the clinical reference had *R*^2^ 0.129 and MAE 6.533; the full model had *R*^2^ 0.134 and MAE 6.445. Observation-process variables alone retained a small association (Δ*R*^2^ 0.009, 95% CI 0.001-0.016), whereas dated source values did not. This is compatible with protocol or acquisition-era information and is not a biological effect.

| Endpoint sensitivity | Evaluation | Full minus clinical $\Delta R^2$ | Delta MAE | Delta median AE |
| --- | --- | --- | --- | --- |
| Primary raw endpoint | Participant CV | 0.0154 (0.0056, 0.0253) | -0.060 (-0.101, -0.018) | -0.036 (-0.130, 0.086) |
| Primary raw endpoint | Enrollment-era test | 0.0049 (-0.0205, 0.0308) | -0.091 (-0.206, 0.020) | -0.132 (-0.531, 0.200) |
| Fold-wise 1–99% winsorized refit | Participant CV | 0.0157 (0.0060, 0.0258) | -0.061 (-0.099, -0.021) | -0.024 (-0.125, 0.080) |
| Earlier-era 1–99% winsorized refit | Enrollment-era test | 0.0084 (-0.0151, 0.0328) | -0.100 (-0.206, 0.004) | -0.142 (-0.533, 0.182) |

Winsorization limits were learned only in each training partition; every preprocessing and ridge step was refitted. The internal information increment persisted, but training-defined endpoint bounding did not establish an enrollment-era advantage. This sensitivity used the enrollment-era split, not the stricter pre-2020-outcome training design. It did not replace the primary endpoint or the separate strict-calendar analysis.

#### S6. Medication-state and subgroup sensitivities

| Sensitivity population | Cutoffs | Participants | $\Delta R^2$ (95% CI) | Delta MAE (95% CI) |
| --- | --- | --- | --- | --- |
| Exam-state-adjusted full cohort | 5,404 | 1,058 | 0.019 (0.009, 0.029) | -0.069 (-0.113, -0.027) |
| Same recorded exam state | 1,511 | 537 | 0.018 (0.003, 0.034) | -0.053 (-0.131, 0.025) |
| ON to ON | 1,348 | 470 | 0.017 (0.001, 0.034) | -0.039 (-0.122, 0.044) |
| OFF to OFF | 163 | 108 | 0.029 (-0.015, 0.076) | -0.168 (-0.370, 0.024) |
| Known state switched | 388 | 247 | -0.000 (-0.026, 0.027) | -0.084 (-0.219, 0.046) |
| LEDD history observed | 4,918 | 948 | 0.019 (0.009, 0.030) | -0.071 (-0.116, -0.027) |

These are observational sensitivities, not medication-effect estimates. The full-minus-clinical internal Δ*R*^2^ was 0.022 (0.005-0.039) in female and 0.012 (0.000-0.025) in male participants. By age it was 0.028 (0.010-0.048) below 60 years, 0.009 (-0.003 to 0.021) at 60-69 years and 0.006 (-0.014 to 0.025) at 70 years or older. Every corresponding temporal-holdout interval included zero. These descriptive summaries neither test interactions nor establish subgroup fairness.

#### S7. Evidence composition and learned-model negative controls

Here sMAE denotes standardized mean absolute error: errors are divided by each target’s fold-local training sample standard deviation, averaged within target and then equally across targets in a source. All models use the same reference scale; the main Methods specify the fitting partitions.

| Representation | MAE | $R^2$ | Calibration slope |
| --- | --- | --- | --- |
| Clinical trajectory and context | 6.766 | 0.102 | 0.958 |
| Hard dominant evidence | 6.764 | 0.103 | 0.955 |
| Continuous evidence composition | 6.726 | 0.114 | 0.917 |
| Raw source dynamics | 6.706 | 0.118 | 0.936 |
| Raw dynamics plus evidence | 6.716 | 0.117 | 0.900 |

Continuous evidence improved on hard assignment (Δ*R*^2^ 0.011, 95% CI 0.002-0.019; delta MAE -0.038, -0.076 to -0.000), but adding evidence coordinates after raw source dynamics changed Δ*R*^2^ by -0.000 (-0.006 to 0.005) and MAE by 0.010 (-0.016 to 0.037). The coordinates improve interpretability rather than predictive information once raw dated values are present.

| Masked source | Clinical ridge sMAE | All-source ridge sMAE | Learned evidence-state sMAE | Learned 90% coverage |
| --- | --- | --- | --- | --- |
| Cognition | 0.694 | 0.701 | 0.714 | 0.902 |
| DaT-SPECT | 0.642 | 0.645 | 0.655 | 0.901 |
| Molecular | 0.661 | 0.664 | 0.682 | 0.892 |

The learned partial-view model reconstructed every masked source worse than the clinical ridge reference. Three complete internal neural runs achieved *R*^2^ 0.125, 0.131 and 0.124, but their increments over the transparent baseline were not securely supported. Removing molecular input from the primary neural run improved *R*^2^ by 0.005 (full-minus-ablated 95% CI -0.008 to -0.001), providing no evidence of useful molecular dependence.

Across 50 fixed initializations trained in the earlier era and tested in 442 post-2020 participants, *R*^2^ ranged from 0.082 to 0.169 (mean 0.145, SD 0.021) and MAE from 6.354 to 6.716 (mean 6.471, SD 0.087). Thirty-six runs exceeded the full ridge *R*^2^ of 0.136, but only 24 improved its MAE of 6.451. No seed was selected, and the neural candidate was not promoted.

#### S8. Participant-disjoint selective-uncertainty results

All uncertainty representations shared the same full source-time mean predictor. Lower area under the risk–coverage curve (AURC) is better.

| Representation | AURC 50%-100% | MAE at 90% | MAE at 80% | MAE at 70% | MAE at 50% |
| --- | --- | --- | --- | --- | --- |
| Clinical uncertainty | 6.465 | 6.637 | 6.526 | 6.391 | 6.207 |
| Generic multimodal uncertainty | 6.449 | 6.621 | 6.543 | 6.375 | 6.198 |
| Source-aware uncertainty | 6.437 | 6.614 | 6.478 | 6.396 | 6.129 |

The source-aware-minus-clinical AURC difference was -0.027 (95% participant-bootstrap CI -0.092 to 0.036). The point estimate was -0.030 under site-disjoint evaluation and reversed to 0.054 under strict calendar evaluation.

| Evaluation and representation | Row coverage | Simultaneous participant coverage | Mean width | Error Spearman rho |
| --- | --- | --- | --- | --- |
| Participant CV, clinical | 0.979 | 0.909 | 49.240 | 0.109 |
| Participant CV, generic multimodal | 0.979 | 0.909 | 48.647 | 0.115 |
| Participant CV, source-aware | 0.977 | 0.906 | 47.820 | 0.115 |
| Site-disjoint, source-aware | 0.985 | 0.927 | 53.874 | 0.108 |
| Strict calendar, source-aware | 0.980 | 0.966 | 48.294 | 0.078 |

The high row coverage is a consequence of calibrating to the maximum error within each participant. It must not be read as a 97.7% nominal row-wise interval. The intended 90% target was simultaneous participant coverage, which was 90.6% internally. Source-aware uncertainty did not demonstrate better selective discrimination and reversed across the calendar boundary; the locked result was ABSTAIN.

Availability strata were highly imbalanced: 5,245 of 5,404 cutoffs had both DaT-SPECT and MoCA, 150 had MoCA only, 7 had DaT-SPECT only and 2 had neither. Rare patterns cannot support standalone coverage claims.

#### S9. Semisynthetic validity-gate results

| Condition | Median internal delta AURC | Median calendar delta AURC | Participant coverage | Overlap detected | Naive promotion, % (95% CI) | Final promotion, % (95% CI) |
| --- | --- | --- | --- | --- | --- | --- |
| Null source effect | 0.033 | 0.031 | 0.907 | 0.000 | 0.80<br>(0.49-1.30) | 0.35<br>(0.17-0.72) |
| Stable source effect | -0.363 | -0.353 | 0.907 | 0.000 | 69.40<br>(67.34-71.38) | 68.60<br>(66.53-70.60) |
| Participant leakage | -1.506 | -1.506 | 0.904 | 1.000 | 94.70<br>(93.63-95.60) | 0.00<br>(0.00-0.19) |
| Calendar reversal | -0.361 | 0.155 | 0.907 | 0.000 | 71.10<br>(69.07-73.04) | 5.25<br>(4.36-6.32) |

The complete gate reduced null promotion to 0.35%, detected every constructed participant overlap and blocked final promotion in 94.75% of calendar-reversal experiments while retaining 68.6% stable-effect power. The residual 5.25% calendar-shift promotion rate is deliberately visible: the gate is a finite-sample safeguard, not an oracle. These results validate the implemented decision logic under controlled failure modes, not PD biology or clinical utility.

#### S10. PDBP transport and diagnosis-modality intersection

| Model | n | MAE | $R^2$ | Calibration slope |
| --- | --- | --- | --- | --- |
| PPMI participant-held-out | 1,256 | 6.428 | 0.134 | 0.997 |
| PDBP raw PPMI transport | 156 | 6.169 | -0.077 | 0.321 |
| PDBP recalibrated PPMI transport | 156 | 5.882 | 0.005 | 0.741 |
| PDBP calibration-mean reference | 156 | 5.860 | -0.009 | Not defined |
| PDBP calibration-trained ridge | 156 | 5.927 | -0.031 | 0.241 |

Recalibrated transport minus the PDBP calibration-mean reference had delta MAE 0.022 (95% CI -0.202 to 0.245). Although its calibration slope entered the prespecified range, the no-material-MAE-degradation condition failed.

| PDBP image or cohort audit item | Result |
| --- | --- |
| Outer bundle size | 3.49 GiB |
| Members indexed | 4,917 |
| Nested ZIP / direct DCM / direct IMA members | 1,230 / 876 / 51 |
| SPECT catalog rows / unique references | 2,457 / 2,442 |
| Catalog references present | 2,442 of 2,442 |
| DaTSCAN rows / unique references present | 1,045 / 1,030 of 1,030 |
| DaTSCAN participants | 139 |
| DaTSCAN participants with any raw MDS-UPDRS | 138 |
| DaTSCAN participants with a complete 11-pair examination | 137 |
| DaTSCAN participants with complete baseline and 9–15-month examinations | 78 |
| DaTSCAN participants with exact idiopathic-PD diagnosis | 0 |
| DaTSCAN participants in diagnosis-change form | 53 |
| DaTSCAN participants with PD in initial or recent change diagnosis | 0 |
| Eligible participants for multimodal PD imaging transport | 0 |

The archive files and side-specific motor trajectories were present. The failure occurred at the diagnosis–modality–time intersection: the local DaTSCAN subset was dominated by dementia with Lewy bodies and related cognitive strata and supplied no exact idiopathic-PD imaging cohort with the required timed outcome. No PDBP image was opened, no SBR was estimated and no multimodal imaging-transport claim was made.

#### S11. Actual records with similar clinical histories and different evidence support

Cases P1-A and P1-B were selected from thirty pairs meeting the baseline-only display rules in Methods. This descriptive analysis was added after the primary evaluation; it did not change the models, folds or outcomes. The cases share a training-fold reference, so their relative evidence scores are comparable. They are not medication-matched and do not identify the cause of their different observed trajectories.

**Table 16:** Measurements available at the selected cutoffs. Ages are months since the source observation, not participant age. An unavailable measurement is not a normal result.

| Measurement | P1-A | P1-B |
| --- | --- | --- |
| MDS-UPDRS Part III total | 10 | 11 |
| Prior motor slope, points/year | 1.3 | 1.5 |
| Prior visits / history span, months | 3 / 17 | 4 / 16 |
| Participant age, years | 72.3 | 71.3 |
| Diagnosis-derived duration, years | 1.7 | 2.5 |
| Mean putamen SBR / source age | 0.835 / 0 | Not observed |
| MoCA / source age | 19 / 0 | 25 / 0 |
| NfL source age | Not observed | 0 |
| GCase source age | Not observed | 14 |
| Active LEDD, mg/day | Not recorded | 1,300 |

**Table 17:**
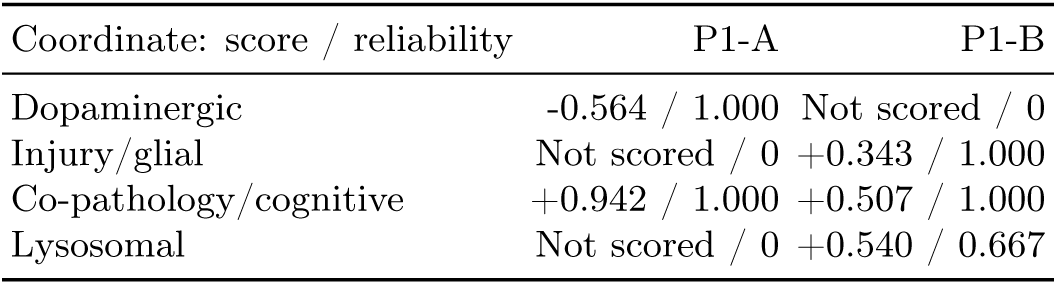
Training-fold-relative evidence score and reliability, reported separately. Score zero denotes the reference midpoint, not absence of disease; reliability summarizes available age-weighted support and is not a calibrated confidence probability.

P1-A’s negative dopaminergic coordinate reflects its position relative to the training PD distribution; it does not imply healthy striatal function. Both records’ co-pathology/cognitive coordinate is supported by MoCA, not proof of a specific co-pathology. P1-B’s injury/glial and lysosomal coordinates are supported by observed NfL and GCase, respectively; these proxies do not establish inflammation or a GBA mutation. With one available source, normalized age weighting leaves the coordinate value unchanged while its reliability declines. The fourteen-month-old GCase example makes this distinction explicit.

**Table 18:** Saved held-out forecasts, in annualized MDS-UPDRS Part III points/year. The representation comparison and nested uncertainty model are separate analyses; their predictions are not successive stages of the primary model.

| Model or observation | P1-A | P1-B |
| --- | --- | --- |
| Clinical trajectory plus context | 4.4 | 1.8 |
| Clinical plus dated source values | 7.5 | 2.8 |
| Clinical plus observation process | 4.0 | 2.2 |
| Full source-time history | 7.3 | 2.5 |
| Hard dominant evidence | 4.5 | 0.4 |
| Continuous evidence composition | 5.9 | 1.6 |
| Separate nested uncertainty mean | 9.1 | 3.0 |
| Observed annualized change | 16.0 | 8.0 |

The actual follow-up examinations occurred at nine months for P1-A (motor total 22) and twelve months for P1-B (total 19). Annualization uses each observed interval, rather than relocating either examination to month twelve. The main article’s patient-evidence figure displays only the nested uncertainty model’s interval around its own mean; it is not an interval for the full source-time or evidence-composition prediction. These records illustrate how measurements, support and forecasts differ, not validated individual benefit or a reason to change treatment.

#### S12. Examination provenance and regional interpretation

The main figures distinguish three levels: observed examination findings, regional imaging evidence, and model-derived summaries. Original records were reconciled after outcome-blind case selection; this descriptive audit did not alter the primary monthly-median spine, refit a model or select a more favorable example.

| Case | Examination state | Left sum | Right sum | Signed ratio | Total III |
| --- | --- | --- | --- | --- | --- |
| P1-A | Unknown | 4 | 0 | -1.000 | 10 |
| P1-B | OFF | 0 | 8 | 1.000 | 11 |
| P1-B | ON | 1 | 7 | 0.750 | 11 |

The P1-B monthly motor total is 11 and its median signed ratio is 0.875. Numeric medians can combine states, and the source examinations must not be interpreted as a controlled medication challenge. In P1-A, a ratio at the -1 boundary comes from four left-side paired points and zero right-side points. Total motor burden is 10, including non-paired items; extremity of the ratio is not extremity of global severity. Gait/freezing items are 1/0 in P1-A and 0/0 in both P1-B examinations. These limited items do not establish a formal gait or motor subtype.

P1-A’s original scan row has left/right putamen SBR 0.96/0.71 and caudate SBR 2.45/1.89, both in the cutoff month. Putamen mean is 0.835 and signed ratio is -0.1497; caudate mean is 2.170 and signed ratio is -0.1290. The caudate values are a source reconciliation, not added primary predictors. No observed DaT-SPECT value is available for P1-B in the frozen rolling record. We do not infer a hemisphere from its motor signs alone.

The regional pathway visualization appears earlier in Figure 7. The anatomical routes provide context for regional imaging, informed by human basal-ganglia connectivity work [18]. They omit indirect and hyperdirect pathways and do not encode excitation, inhibition or connection strength. P1-A’s lower right putamen binding supports a spatially specific imaging description; MoCA, NfL and GCase remain non-localizing cognitive or biomarker descriptions. Adding a detailed reference brain does not increase the spatial resolution of those measurements.

